# Body mass index and subfertility: multivariable regression and Mendelian randomization analyses in the Norwegian Mother, Father and Child Cohort Study

**DOI:** 10.1101/2021.06.21.21259264

**Authors:** Álvaro Hernáez, Tormod Rogne, Karoline H. Skåra, Siri E. Håberg, Christian M. Page, Abigail Fraser, Stephen Burgess, Deborah A. Lawlor, Maria Christine Magnus

## Abstract

**Background:** Higher body mass index (BMI) is associated with subfertility in women and men. This relationship is further substantiated by a few small randomized-controlled trials of weight reduction and success of assisted reproduction. The aim of the current study was to expand the current evidence-base by investigating the association between BMI and subfertility in men and women using multivariable regression and Mendelian randomization.

**Methods and findings:** We studied 34,157 women (average age 30, average BMI 23.1 kg/m^2^) and 31,496 men (average age 33, average BMI 25.4 kg/m^2^) who were genotyped and are participating in the Norwegian Mother, Father and Child Cohort Study. Self-reported information was available on time-to-pregnancy and BMI. A total of 10% of couples were subfertile (time-to-pregnancy ≥12 months). Our findings support a J-shaped association between BMI and subfertility in both sexes using multivariable logistic regression models. Non-linear Mendelian randomization validated this relationship. A 1 kg/m^2^ greater genetically predicted BMI was linked to 15% greater odds of subfertility (95% confidence interval 4% to 28%) in obese women (≥30.0 kg/m^2^) and 14% lower odds of subfertility (−25% to -3%) in women with BMI <20.0 kg/m^2^. A 1 kg/m^2^ higher genetically predicted BMI was linked to 23% greater odds of subfertility (6% to 43%) among obese men and 36% decreased odds (−62% to 7%) among men BMI <20.0 kg/m^2^. A genetically predicted BMI of 23 and 25 kg/m^2^ was linked to the lowest subfertility risk in women and men, respectively. The main limitations of our study were that we did not know whether the subfertility was driven by the woman, man, or both; the exclusive consideration of individuals of northern European ancestry; and the limited amount of participants with obesity or BMI values <20.0 kg/m^2^.

**Conclusions:** We observed a J-shaped relationship between BMI and subfertility in both sexes, when using both a standard multivariable regression and Mendelian randomization analysis, further supporting a potential causal role of BMI on subfertility.

**AUTHOR SUMMARY:** *WHY WAS THIS STUDY DONE?:*

- Higher body mass index (BMI) in both women and men is associated with subfertility in in observational studies. A few small randomized-controlled trials of weight reduction have reported an increased success of assisted reproduction in women. In addition, women with BMI <18.5 kg/m^2^ have lower conception rates with assisted reproduction technologies. A non-linear relationship between BMI and subfertility is suggested.
- We aimed to investigate the association between BMI and subfertility using both a standard multivariable regression and a Mendelian randomization approach.

*WHAT DID THE RESEARCHERS DO AND FIND?:*

- We examined the relationship between BMI and subfertility (time-to-pregnancy ≥12 months) among all men and women in the Norwegian Mother, Father and Child Cohort Study with available genotype information and anthropometric data (34,157 women, 31,496 men).
- We observed a J-shaped relationship between BMI and subfertility in both sexes, when using both a standard multivariable regression and Mendelian randomization analysis.

*WHAT DO THESE FINDINGS MEAN?:*

- Together with previous observational and trial evidence, findings support a causal effect of overweight/obesity on subfertility in women and men.
- Our findings expand the current evidence by also indicating that individuals at the lower end of the BMI distribution (<20 kg/m^2^) may have an increased risk of subfertility.
- Current advice and support for overweight and obese subfertile couples to lose weight should continue. Additionally, clinicians should consider appropriate advice to those with low BMI on whether they should increase weight to obtain a normal BMI.

## INTRODUCTION

Body weight is associated with the ability to reproduce [1, 2]. In observational studies, high body mass index (BMI) in women is associated with greater risk of subfertility, commonly defined as trying to conceive without success for ≥12 months [3, 4], or a lower success of assisted reproductive technology (ART) [5]. In addition, women with BMI <18.5 kg/m^2^ have a lower chance of ART success [6], supporting the hypothesis of a non-linear relationship between BMI and subfertility. In men, subfertility appears to be more prevalent among those who are overweight or obese, and obese men are also overrepresented among ART users [7, 8]. There is some experimental evidence indicating that weight reduction interventions among women improve their likelihood of success after undergoing an ART treatment [9].

BMI is closely linked to a broad range of other characteristics which are also related to subfertility [10, 11]. In addition, previous studies addressing the role of BMI on subfertility have not accounted for the partner’s BMI, which may add to the effect as individuals with greater BMI values are more likely to have a partner with elevated BMI (assortative mating) [12]. Thus, it remains unclear whether there is a causal relationship between BMI and subfertility, or if the association is due to confounding or partner’s BMI. The use of complementary methodological approaches could contribute to a better understanding of this matter. Mendelian randomization (MR) uses genetic variants that are robustly related to an exposure (e.g. BMI) to retrieve the unconfounded effect of that exposure on an outcome (e.g. subfertility) [13]. Results from MR are less likely to be confounded by the socioeconomic and behavioral factors that commonly affect conventional regression analyses but, at the same time, are susceptible to bias due to weak instruments and horizontal pleiotropy [14]. Given the different sources of bias between multivariable regression and MR, when findings agree, it increases confidence in the consistent results reflecting a causal effect [15].

Our aim was to investigate the association between BMI and subfertility in women and men using multivariable logistic regression and MR.

## METHODS

### The Norwegian Mother, Father and Child Cohort Study

Our study included participants in the Mother, Father and Child Cohort Study (MoBa) [16, 17]. The MoBa Study is a population-based pregnancy cohort study conducted by the Folkehelseinstituttet/Norwegian Institute of Public Health. Participants were recruited from all over Norway from 1999-2008. The women consented to participation in 41% of the pregnancies. The cohort now includes 114,500 children, 95,200 mothers and 75,200 fathers. The current study is based on version #12 of the quality-assured data. The establishment of MoBa and initial data collection was based on a license from the Norwegian Data Protection Agency and approval from The Regional Committees for Medical and Health Research Ethics. The MoBa cohort is now based on regulations related to the Norwegian Health Registry Act.

For the current study, we defined a subsample of parents with available genotype data and pre-pregnancy information on BMI. The genotype data used in this study come from blood samples obtained from both parents during pregnancy [18] and followed the pipeline described by Helgeland et al regarding genotype calling, imputation, and quality control [19]. We have described our work according to the STROBE guidelines for reporting MR (**S1 Checklist**) and cohort studies (**S2 Checklist**).

### BMI

Maternal and paternal pre-pregnancy weight and height were reported in the questionnaire completed at recruitment and used to calculate BMI as weight in kilograms divided by the squared height in meters. Extreme BMI values <9 or >90 kg/m^2^ were excluded.

### Genetic risk score for BMI

We used the results from the most recent genome-wide association study (GWAS) of BMI to create the genetic instrument in our analysis [20]. This GWAS included approximately 700,000 individuals of European ancestry (none of them participated in the MoBa cohort) that yielded 941 independent single nucleotide polymorphisms (SNPs) associated with BMI [20]. 896 of the 941 SNPs were available in the MoBa genotype data. We computed a weighted genetic risk score (GRS) by multiplying the number of risk alleles by the effect estimate of each variant and dividing by the total number of SNPs [21].

### Subfertility

At the time of recruitment, women were asked whether the pregnancy was planned, and to provide information on how many months it had taken them to conceive [17]. The answer options were less than one month, 1-2 months, and 3 or more months. If the mother had used ≥3 months, she was asked to further specify exactly how many months the couple had been trying to conceive. Subfertility was defined as time-to-pregnancy ≥12 months or having used ART. Those reporting a time-to-pregnancy <12 months or involved in unplanned pregnancies were included in the reference group.

### Other variables

From the MoBa questionnaires, we gathered information on age (continuous), educational level (years of education equivalent to the US system [22, 23], continuous), cigarette smoking (never smokers, former smokers, having quitted smoking by 12^th^ –mothers– or 18^th^ gestational week –fathers–, or being a current smoker), and previous number of deliveries (0, 1, 2, or ≥3).

### Ethical approval

The data collection in MoBa is approved by the Norwegian Data Inspectorate. Participants provided a written informed consent before joining the cohort. This project was approved by the Regional Committee for Medical and Health Research Ethics of South/East Norway (reference: 2017/1362).

### Statistical analyses

We used means and standard deviations to describe normally distributed continuous variables, medians and 1^st^-3^rd^ quartiles for non-normally distributed continuous variables, and proportions for categorical variables. We assessed differences in baseline characteristics among subfertile and non-subfertile parents using t-tests for normally distributed continuous variables, Mann-Whitney U-tests for non-normally distributed continuous variables, and chi-squared tests in categorical variables.

We first evaluated the presence of a linear relationship between BMI and subfertility in women and men separately by standard logistic regressions. We examined the evidence for a non-linear association by assessing the relationship between a 1 kg/m^2^ increase in measured BMI and subfertility odds in BMI categories defined by current WHO guidelines: underweight and normal-low weight (<20.0 kg/m^2^), normal weight (20.0-24.9 kg/m^2^), overweight (25.0-29.9 kg/m^2^), and obesity (≥30.0 kg/m^2^). We also assessed whether a model using smoothed cubic splines (*K*+4 degrees of freedom) to model the relationship between BMI and subfertility fitted the data better than a simple linear term using a likelihood ratio test. All logistic regression models were adjusted for age, education years, smoking, and number of previous deliveries. We computed clustered standard errors to account for dependency between women/men who participated with more than one pregnancy.

In the MR analyses, we used a linear regression model to obtain a genetically predicted BMI using the GRS for BMI as a predictor. We assessed the linear relationship between genetically predicted BMI and subfertility by logistic regression models. We explored non-linear associations by investigating the association between a 1 kg/m^2^ increase in the genetically predicted BMI and subfertility within residual BMI categories using WHO definitions as previously described. Residual BMI is defined as the participant’s reported BMI minus the genetically predicted BMI. The stratification according to residual BMI allows the comparison of participants who would have a similar BMI if they had the same genetic information and is a strategy to minimize collider bias [24].

We also applied a fractional polynomial method to calculate non-linear MR estimates of BMI on subfertility odds. In this procedure, we first divided the population into 100 strata of equal number of participants according to the residual BMI. We then calculated the linear MR estimate in each stratum (the association of the GRS with the outcome divided by the association of the GRS with the exposure). Finally, we performed a meta-regression of these estimates against the mean value of the reported BMI in each of the 100 strata using a fractional polynomial model as previously described [24, 25]. We also calculated a fractional polynomial test, which assessed if the model using fractional polynomials to model the relationship between genetically predicted BMI and subfertility fitted the causal effect estimates better than a model with a simple linear term.

Three assumptions must be met in a valid MR study: the genetic instrument is robustly associated with the exposure, the genetic instrument is only linked to the outcome through the exposure of interest, and there is no confounding of the genetic instrument-outcome associations [26]. The strength of the genetic instrument (the association between the GRSs and BMI) was assessed in women and men separately using linear regressions, *F*-statistics, and R^2^ coefficients of determination. Regarding the second assumption, a common cause of violation is horizontal pleiotropy (i.e. genetic instrumental variables influence other risk factors for the outcome in addition to the exposure of interest) [14]. To check this bias, we assessed the associations between quartiles of the GRS and predefined risk factors for subfertility (age, educational levels, smoking, and number of previous pregnancies). Whenever we found indication of pleiotropic effects, we performed: 1) multivariable MR analyses if a valid genetic instrument could be calculated, i.e. if there were GWAS or meta-analyses of GWAS whose summary data were available [27]; or 2) stratified analyses. We identified summary GWAS data that enabled us to conduct multivariable MR analyses for educational level and smoking initiation and conducted stratified analyses according to age (below vs. over the median). For the multivariable MR accounting for educational level we used the results from the most recent GWAS of education, which included approximately 1.1 million individuals and reported 1,271 independent SNPs [28]. We estimated the genetically predicted years of education using a GRS based on the 1,159 available SNPs in the MoBa genotype data. For the multivariable MR accounting for smoking, we used the summary results of the most recent GWAS, which included more than 1.2 million participants and reported 378 SNPs associated with smoking initiation [29]. In this case, we estimated the genetically determined risk of starting to smoke by a GRS based on the 355 available SNPs in the MoBa genotype data. In both multivariable MR analyses, we estimated the genetically predicted BMI values also including the GRS for education and the GRS for smoking initiation. Similarly, the genetically predicted number of educational years and likelihood of starting to smoke were estimated considering the GRS for BMI in addition to the GRS for the covariate of interest. Finally, we assessed the association between the genetically predicted BMI and subfertility as previously described using models further adjusted for the genetically predicted education years and likelihood of starting to smoke. We further explored unbalanced horizontal pleiotropy by methods developed for use in two sample MR [30-32]. We first generated summary results of the association of each of the 896 SNPs related to BMI with subfertility in a GWAS in the MoBa cohort (full details of these analyses are provided in the Supplemental Materials), and combined them with summary data of their association with BMI [20] to create a two sample MR framework. We performed the two sample MR by different methods, including random effects inverse variance weighted regression, MR-Egger, weighted median method, and MR weighted mode estimator. We checked the presence of horizontal pleiotropy by: estimating the MR-Egger intercept (a deviation from zero would suggest horizontal pleiotropy); comparing the causal estimates obtained in the inverse variance weighted regression, the MR-Egger, and the weighted median methods (a divergence among them would also suggest horizontal pleiotropy); and generating a scatterplot as a visual check for potentially pleiotropic outliers in the variant-specific causal estimates [30-32]. We also estimated between SNP heterogeneity (by the Cochran’s *Q* and the Rücker’s *Q’* statistics according to the inverse variance weighted regression and MR-Egger methods, respectively). Finally, regarding the third MR assumption (lack of confounding of the genetic instrument-outcome associations), all the one sample MR analyses were adjusted for 10 ancestry-informative principal components to account for population stratification [33].

As additional sensitivity analyses: 1) we restricted the analysis to parents reporting having planned their pregnancies (28,328 women –83.0%– and 26,252 men –83.4%–); and 2) we removed the conceptions by ART from the case group (716 and 680 in women and men, respectively –21% of the overall subfertile cases–).

All analyses were performed in R Software version 4.0.3 (packages: *compareGroups, estimatr, ggplot2, miceadds*, and *TwoSampleMR*). Code for data management and statistical analysis is available here: https://github.com/alvarohernaez/MR_BMI_subfertility_MoBa/blob/main/syntax.

## RESULTS

### Study population

Our study population consisted of 34,157 women (30 years old on average, mean pre-pregnancy BMI 23.1 kg/m^2^) and 31,496 men (33 years old on average, mean BMI pre-pregnancy 25.4 kg/m^2^) with singleton pregnancies and information on both BMI and genotype (**Fig 1**). A total of 10% of the couples were subfertile. Women and men who were subfertile were older, had a lower educational level, were more likely to be current/former smokers, and more likely to be trying for a first pregnancy, and had on average greater BMI (**Table 1**).

**Table 1.**
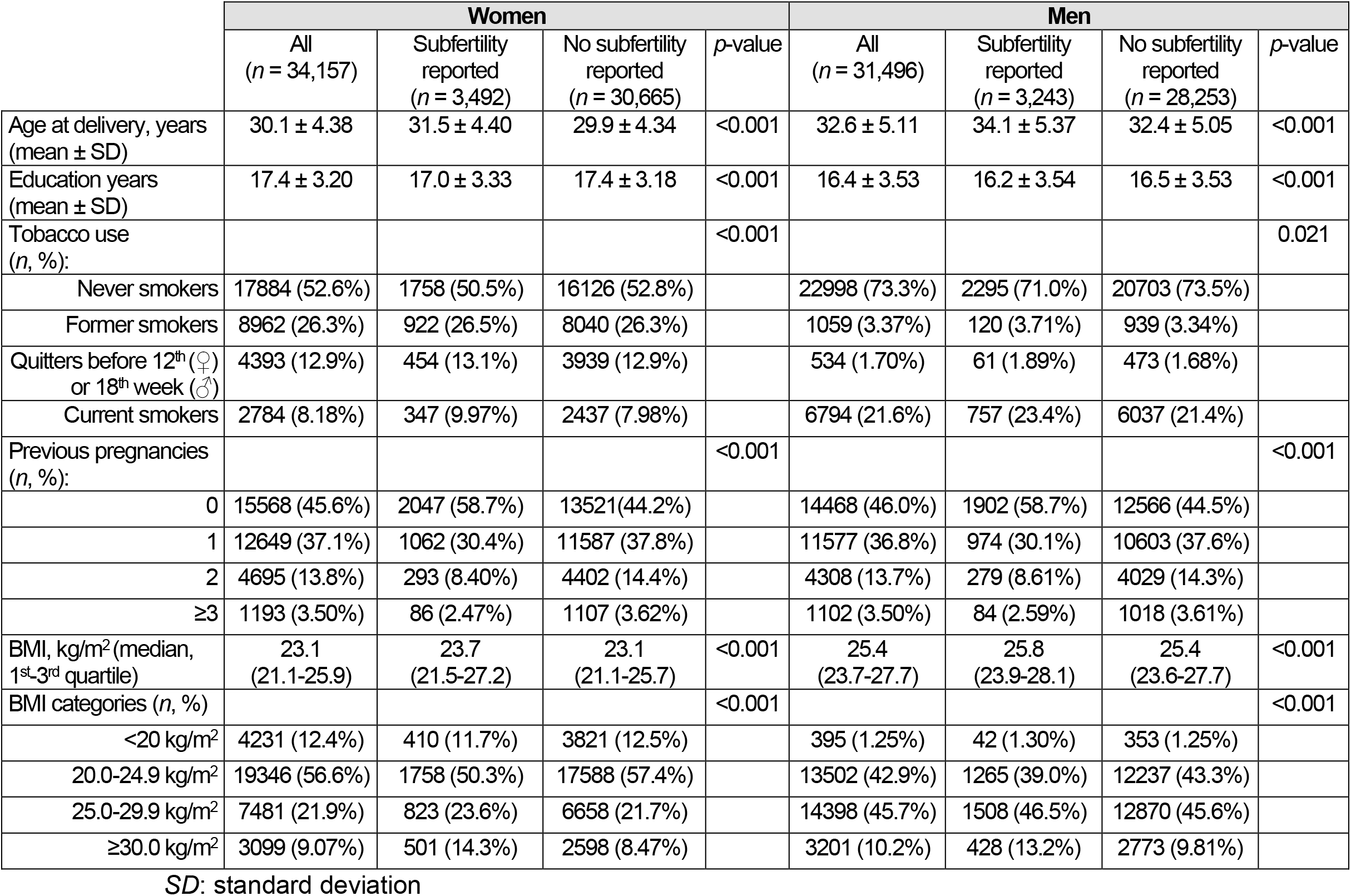
Population characteristics

|  | Women |  |  |  | Men |  |  |  |
| --- | --- | --- | --- | --- | --- | --- | --- | --- |
|  | All<br>(n = 34,157) | Subfertility<br>reported<br>(n = 3,492) | No subfertility<br>reported<br>(n = 30,665) | p-value | All<br>(n = 31,496) | Subfertility<br>reported<br>(n = 3,243) | No subfertility<br>reported<br>(n = 28,253) | p-value |
| Age at delivery, years<br>(mean ± SD) | 30.1 ± 4.38 | 31.5 ± 4.40 | 29.9 ± 4.34 | <0.001 | 32.6 ± 5.11 | 34.1 ± 5.37 | 32.4 ± 5.05 | <0.001 |
| Education years<br>(mean ± SD) | 17.4 ± 3.20 | 17.0 ± 3.33 | 17.4 ± 3.18 | <0.001 | 16.4 ± 3.53 | 16.2 ± 3.54 | 16.5 ± 3.53 | <0.001 |
| Tobacco use<br>(n, %): |  |  |  | <0.001 |  |  |  | 0.021 |
| Never smokers | 17884 (52.6%) | 1758 (50.5%) | 16126 (52.8%) |  | 22998 (73.3%) | 2295 (71.0%) | 20703 (73.5%) |  |
| Former smokers | 8962 (26.3%) | 922 (26.5%) | 8040 (26.3%) |  | 1059 (3.37%) | 120 (3.71%) | 939 (3.34%) |  |
| Quitters before 12<sup>th</sup> (♀)<br>or 18<sup>th</sup> week (♂) | 4393 (12.9%) | 454 (13.1%) | 3939 (12.9%) |  | 534 (1.70%) | 61 (1.89%) | 473 (1.68%) |  |
| Current smokers | 2784 (8.18%) | 347 (9.97%) | 2437 (7.98%) |  | 6794 (21.6%) | 757 (23.4%) | 6037 (21.4%) |  |
| Previous pregnancies<br>(n, %): |  |  |  | <0.001 |  |  |  | <0.001 |
| 0 | 15568 (45.6%) | 2047 (58.7%) | 13521 (44.2%) |  | 14468 (46.0%) | 1902 (58.7%) | 12566 (44.5%) |  |
| 1 | 12649 (37.1%) | 1062 (30.4%) | 11587 (37.8%) |  | 11577 (36.8%) | 974 (30.1%) | 10603 (37.6%) |  |
| 2 | 4695 (13.8%) | 293 (8.40%) | 4402 (14.4%) |  | 4308 (13.7%) | 279 (8.61%) | 4029 (14.3%) |  |
| ≥3 | 1193 (3.50%) | 86 (2.47%) | 1107 (3.62%) |  | 1102 (3.50%) | 84 (2.59%) | 1018 (3.61%) |  |
| BMI, kg/m<sup>2</sup> (median,<br>1<sup>st</sup>-3<sup>rd</sup> quartile) | 23.1<br>(21.1-25.9) | 23.7<br>(21.5-27.2) | 23.1<br>(21.1-25.7) | <0.001 | 25.4<br>(23.7-27.7) | 25.8<br>(23.9-28.1) | 25.4<br>(23.6-27.7) | <0.001 |
| BMI categories (n, %) |  |  |  | <0.001 |  |  |  | <0.001 |
| <20 kg/m<sup>2</sup> | 4231 (12.4%) | 410 (11.7%) | 3821 (12.5%) |  | 395 (1.25%) | 42 (1.30%) | 353 (1.25%) |  |
| 20.0-24.9 kg/m<sup>2</sup> | 19346 (56.6%) | 1758 (50.3%) | 17588 (57.4%) |  | 13502 (42.9%) | 1265 (39.0%) | 12237 (43.3%) |  |
| 25.0-29.9 kg/m<sup>2</sup> | 7481 (21.9%) | 823 (23.6%) | 6658 (21.7%) |  | 14398 (45.7%) | 1508 (46.5%) | 12870 (45.6%) |  |
| ≥30.0 kg/m<sup>2</sup> | 3099 (9.07%) | 501 (14.3%) | 2598 (8.47%) |  | 3201 (10.2%) | 428 (13.2%) | 2773 (9.81%) |  |

SD: standard deviation

**Fig 1.**
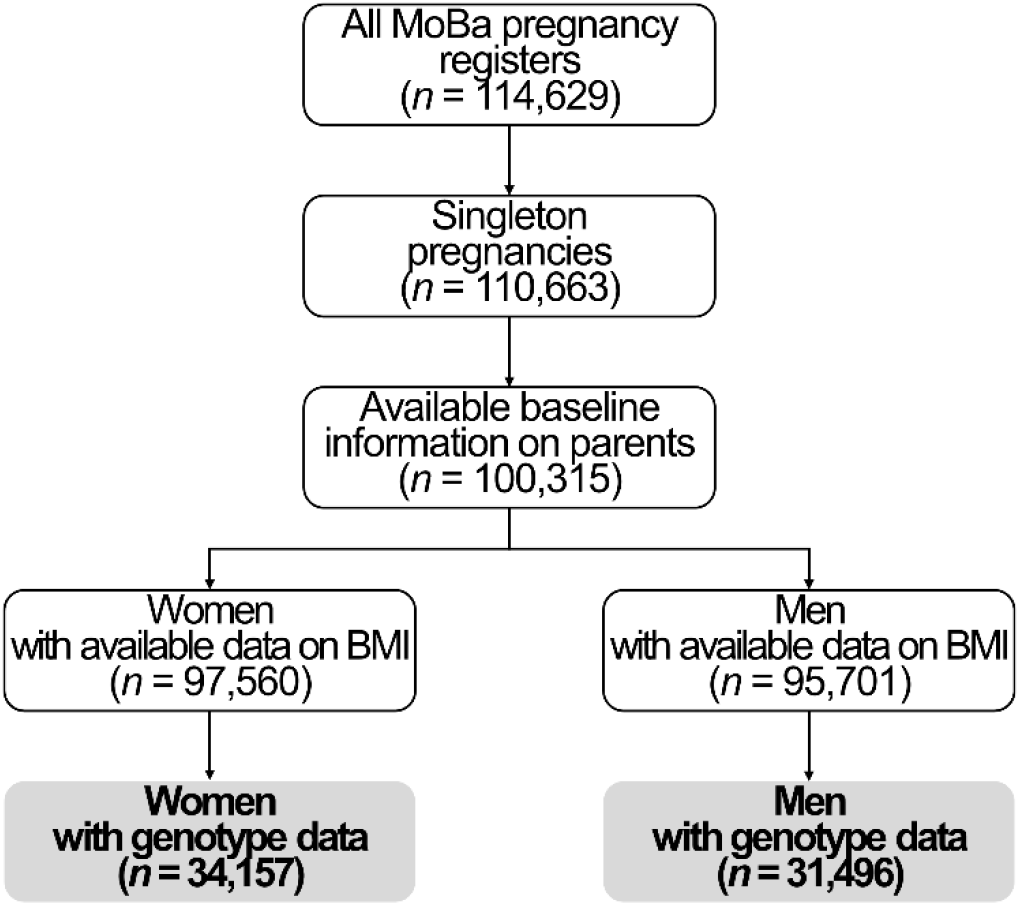
Study flow chart.

### Association between reported BMI and subfertility: multivariable logistic regressions

In the standard multivariable linear association, each 1 kg/m^2^ increase in BMI was linked to 4% greater odds of subfertility in women (odds ratio –OR– 1.04, 95% confidence interval – CI– 1.04 to 1.05, *p* < 0.001) and men (OR 1.04, 95% CI 1.03 to 1.05, *p* < 0.001). However, a non-linear model based on restricted cubic splines fitted the data better than a linear term in both sexes (likelihood ratio tests: *p*_women_ < 0.001, *p*_men_ = 0.035; **Fig 2A** and **2B**). These relationships were J-shaped, with a positive association from BMI values of 22 kg/m^2^ onwards in both sexes. A 1 kg/m^2^ increase in BMI was linked to 4% greater odds of subfertility in women with a BMI between 20.0 and 24.9 kg/m^2^ (OR 1.04, 95% CI 1.00 to 1.08, *p* = 0.031), 10% increased odds in overweight women (OR 1.10, 95% CI 1.04 to 1.16, *p* < 0.001), and 4% greater odds in obese women (OR 1.04, 95% CI 1.01 to 1.07, *p* = 0.005). On the contrary, a 1 kg/m^2^ increment in BMI was associated with 14% lower odds of subfertility in women with BMI <20.0 kg/m^2^ (OR 0.86, 95% CI 0.75 to 0.98, *p* = 0.027) (**Fig. 2A**). In men, a 1 kg/m^2^ increase in BMI was linked to 5% greater odds of subfertility in participants with a BMI between 20.0 and 24.9 kg/m^2^ (OR 1.05, 95% CI 1.00 to 1.10, *p* = 0.068), 6% increased odds in overweight men (OR 1.06, 95% CI 1.02 to 1.10, *p* = 0.007), and 8% greater odds in obese men (OR 1.08, 95% CI 1.03 to 1.12, *p* < 0.001), and there was no evidence of an association in those with BMI values <20.0 kg/m^2^ (OR 0.80, 95% CI 0.51 to 1.25, *p* = 0.322) (**Fig. 2B**).

**Fig 2.**
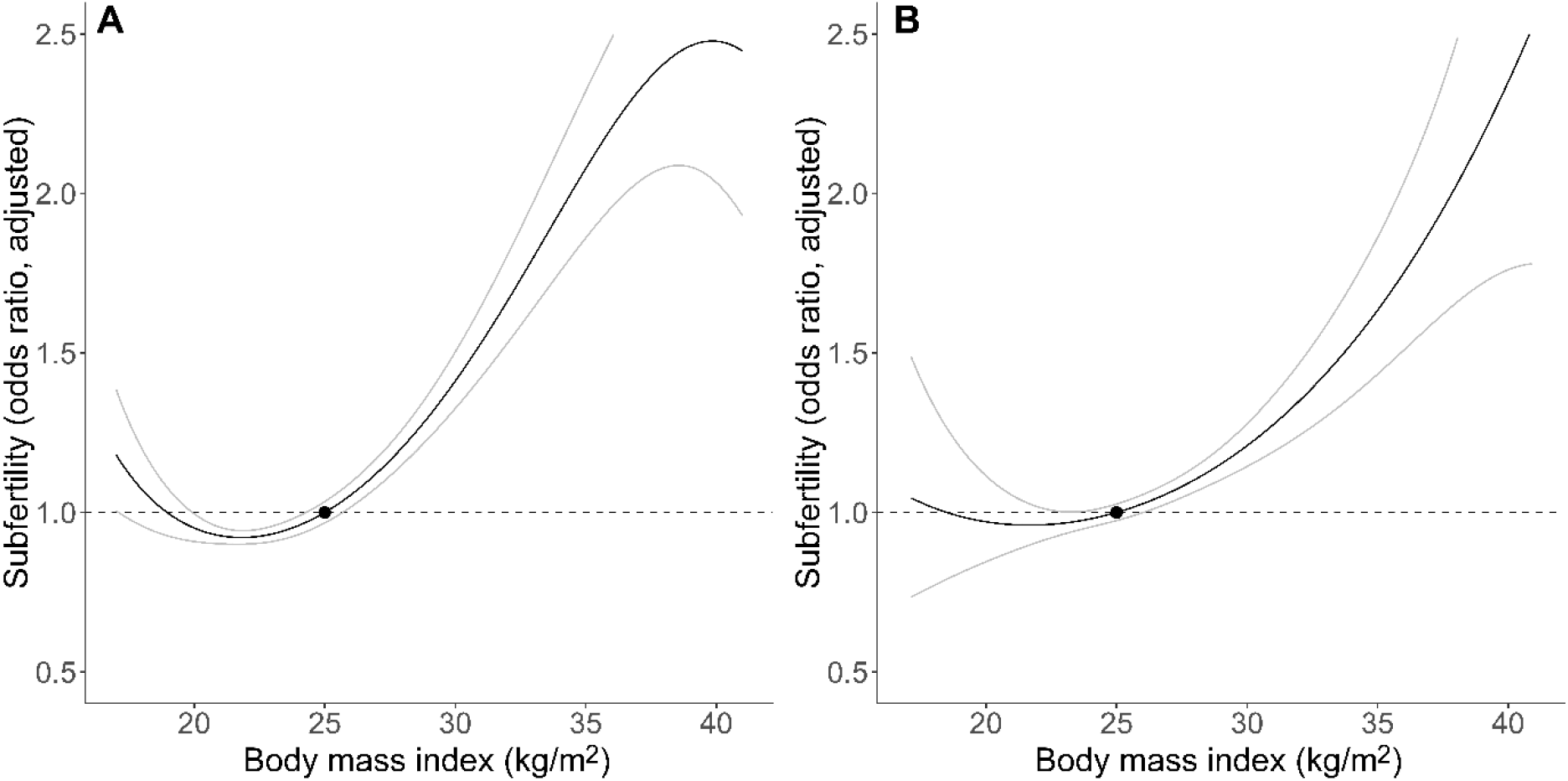
Association between reported body mass index and subfertility in women (A) and men (B). Non-linear logistic regression analyses (smoothed cubic splines) adjusted for age, education level, smoking, and number of previous pregnancies. A BMI of 25 kg/m^2^ was set as reference (black dot). Gray lines represent 95% confidence intervals

### MR analyses on BMI and subfertility in women

Each one unit increase in the GRS was linked to a BMI increase of 0.044 kg/m^2^ (95% CI 0.041 to 0.046, *p* < 0.001, 5.63% of BMI variation explained, *F*-statistic = 1,388). There was evidence of a J-shaped relationship between the genetically predicted BMI and subfertility in women (fractional polynomial test *p*-value for non-linearity = 0.033), which was positive for BMI values ≥ 23.3 kg/m^2^ (**Fig. 3**). A 1 kg/m^2^ increase in genetically predicted BMI was linked to 15% greater odds of subfertility in obese women (OR 1.15, 95% CI 1.04 to 1.28, *p* = 0.010), 14% lower odds in women with BMI <20.0 kg/m^2^ (OR 0.86, 95% CI 0.75 to 0.97, *p* = 0.015), and unrelated to subfertility in those with BMI values between 20.0 and 24.9 kg/m^2^ (OR 0.99, 95% CI 0.94 to 1.04, *p* = 0.645) and in overweight women (OR 1.03, 95% CI 0.96 to 1.11, *p* = 0.397).

**Fig 3.**
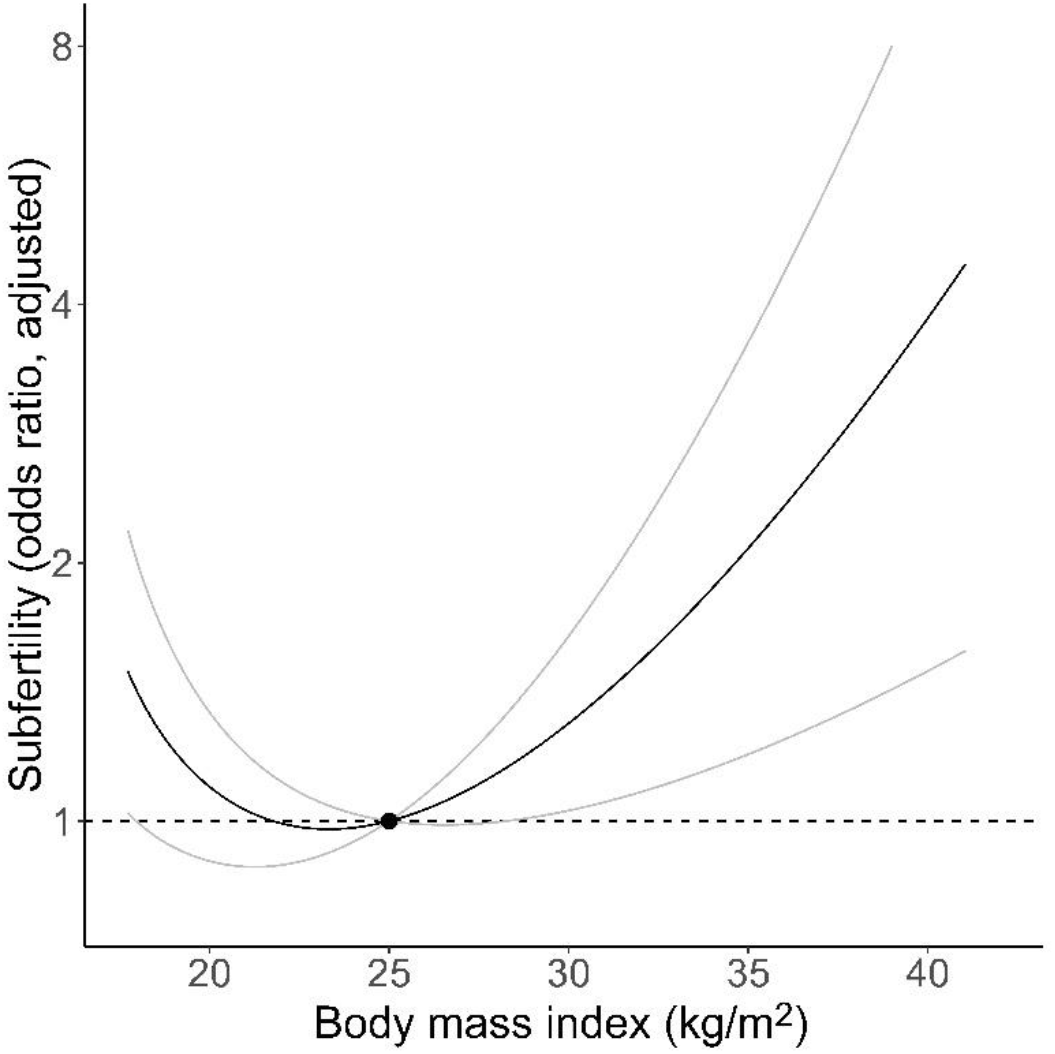
Mendelian randomization analysis of body mass index and subfertility in women. A BMI of 25 kg/m^2^ was set as reference (black dot). Gray lines represent 95% confidence intervals.

**Fig 4.**
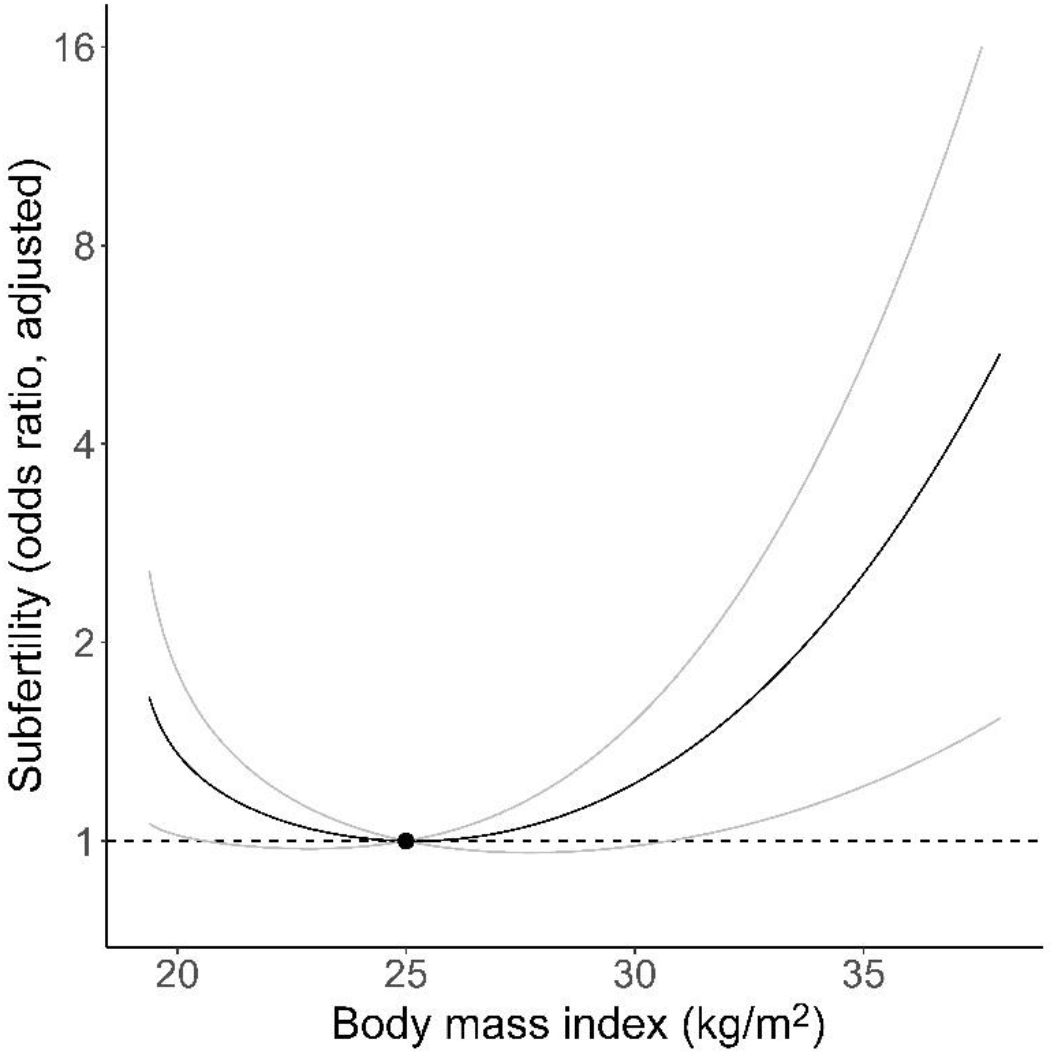
Mendelian randomization analysis of body mass index and subfertility in men. A BMI of 25 kg/m^2^ was set as reference (black dot). Gray lines represent 95% confidence intervals.

### MR analyses on BMI and subfertility in men

Each one unit increase in the GRS was linked to a BMI increase of 0.033 kg/m^2^ in men (95% CI 0.031 to 0.035, *p* < 0.001, 5.01% of BMI variation explained, *F*-statistic = 1,202). We observed a non-linear, J-shaped association between genetically predicted BMI and subfertility in men (*p*-value for non-linearity = 0.018), which was positive for BMI values ≥ 25.2 kg/m^2^ (**Fig. 3**). A 1 kg/m^2^ increment in genetically predicted BMI was linked to 23% greater odds of subfertility in obese men (OR 1.23, 95% CI 1.06 to 1.43, *p* = 0.006), marginally related to 36% lower odds in men with BMI <20.0 kg/m^2^ (OR 0.64, 95% CI 0.38 to 1.07, *p* = 0.090; only 1.25% of all men belonged to this category), and unrelated to subfertility in men with BMI values between 20.0 and 24.9 kg/m^2^ (OR 0.96, 95% CI 0.88 to 1.04, *p* = 0.341) and in overweight participants (OR 1.01, 95% CI 0.93 to 1.09, *p* = 0.894).

### Verification of MR assumptions

Regarding horizontal pleiotropy, we observed an inverse relationship of GRS for BMI with education and age, and there was a lower proportion of never smokers in participants with high GRS values in both women (**S1 Table**) and men (**S2 Table**). In both sexes, we observed similar J-shaped associations between BMI and subfertility in the multivariable MR accounting for education and smoking to those observed in the main analyses (**Table 2**; **S1** and **S2 Figs**). In relation to age, we stratified our analyses in participants below and over the median age (30 years in women, 32 years in men). Genetically predetermined BMI had a similar non-linear, J-shaped associations with subfertility in both age groups as seen in the main analyses (**Table 2, S3 Fig**).

**Table 2.** Multivariable and age-stratified MR analyses

|  | MR: main analyses | MR + education years (multivariable) | MR + smoking initiation (multivariable) | MR: age of delivery < median | MR: age of delivery > median |
| --- | --- | --- | --- | --- | --- |
| **Women** |  |  |  |  |  |
| **Linear MR** |  |  |  |  |  |
| OR for $\Delta 1 \text{ kg/m}^2$ (whole population) | 1.02<br>(0.99 to 1.06) | 1.02<br>(0.98 to 1.06) | 1.02<br>(0.98 to 1.06) | 1.01<br>(0.95 to 1.06) | 1.04<br>(0.99 to 1.09) |
| **Non-linear MR** |  |  |  |  |  |
| Fractional polynomial test (*p*-value for non-linearity) | 0.033 | 0.029 | 0.047 | 0.067 | 0.030 |
| OR for $\Delta 1 \text{ kg/m}^2$ (stratified analyses): | | | | | |
| < 20.0 $\text{kg/m}^2$ | 0.86<br>(0.75 to 0.97) | 0.84<br>(0.73 to 0.97) | 0.85<br>(0.74 to 0.97) | 0.81<br>(0.68 to 0.97) | 0.87<br>(0.72 to 1.04) |
| 20.0-24.9 $\text{kg/m}^2$ | 0.99<br>(0.94 to 1.04) | 0.99<br>(0.94 to 1.04) | 0.98<br>(0.93 to 1.03) | 0.97<br>(0.90 to 1.05) | 1.00<br>(0.94 to 1.07) |
| 25.0-29.9 kg/m<sup>2</sup> | 1.03<br>(0.96 to 1.11) | 1.03<br>(0.96 to 1.12) | 1.04<br>(0.96 to 1.13) | 1.07<br>(0.95 to 1.20) | 1.03<br>(0.93 to 1.13) |
| ≥ 30.0 kg/m<sup>2</sup> | 1.15<br>(1.04 to 1.28) | 1.15<br>(1.03 to 1.29) | 1.16<br>(1.04 to 1.30) | 1.08<br>(0.94 to 1.25) | 1.24<br>(1.06 to 1.46) |
| BMI with lowest subfertility odds | 23.3 kg/m<sup>2</sup> | 23.1 kg/m<sup>2</sup> | 23.6 kg/m<sup>2</sup> | 24.7 kg/m<sup>2</sup> | 22.1 kg/m<sup>2</sup> |
| **Men** |  |  |  |  |  |
| **Linear MR** |  |  |  |  |  |
| OR for Δ1 kg/m<sup>2</sup> (whole population) | 1.01<br>(0.96 to 1.07) | 1.00<br>(0.95 to 1.06) | 1.02<br>(0.98 to 1.09) | 1.06<br>(0.97 to 1.15) | 1.00<br>(0.93 to 1.07) |
| **Non-linear MR** |  |  |  |  |  |
| Fractional polynomial test (*p*-value for non-linearity) | 0.018 | 0.037 | 0.011 | 0.018 | 0.048 |
| OR for Δ1 kg/m<sup>2</sup> (stratified analyses): |  |  |  |  |  |
| < 20.0 kg/m<sup>2</sup> | 0.64<br>(0.38 to 1.07) | 0.73<br>(0.41 to 1.28) | 0.72<br>(0.39 to 1.35) | 0.78<br>(0.39 to 1.55) | 0.73<br>(0.28 to 1.91) |
| 20.0-24.9 kg/m<sup>2</sup> | 0.96<br>(0.88 to 1.04) | 0.95<br>(0.87 to 1.04) | 0.96<br>(0.88 to 1.06) | 0.98<br>(0.86 to 1.11) | 0.94<br>(0.84 to 1.05) |
| 25.0-29.9 kg/m<sup>2</sup> | 1.01<br>(0.93 to 1.09) | 1.00<br>(0.92 to 1.09) | 1.02<br>(0.93 to 1.11) | 1.05<br>(0.92 to 1.19) | 1.00<br>(0.91 to 1.10) |
| ≥ 30.0 kg/m<sup>2</sup> | 1.23<br>(1.06 to 1.43) | 1.24<br>(1.05 to 1.45) | 1.25<br>(1.05 to 1.48) | 1.32<br>(1.06 to 1.64) | 1.19<br>(0.97 to 1.45) |
| BMI with lowest subfertility odds | 25.2 kg/m<sup>2</sup> | 26.7 kg/m<sup>2</sup> | 24.5 kg/m<sup>2</sup> | 24.7 kg/m<sup>2</sup> | 25.5 kg/m<sup>2</sup> |

MR: Mendelian randomization; OR: odds ratio

Further sensitivity analyses using a two sample MR framework indicated no evidence of a linear relationship between BMI and subfertility, no horizontal pleiotropy according to different methods with various assumptions, and no SNP heterogeneity (**S3 Table, S4 Fig**).

### Other sensitivity analyses

Genetically predetermined BMI presented similar non-linear, J-shaped associations with subfertility in both women and men also when restricting the analysis to those with planned pregnancies (**S4 Table, S5 Fig**) and after excluding ART users (**S5 Table, S6 Fig**).

## DISCUSSION

Our findings from multivariable and MR analyses indicate that BMI has a J-shaped association with subfertility in both men and women. Both participants with BMI values <20.0 kg/m^2^ and obese individuals had an increased risk of subfertility. The consistency of the results between multivariable regression and MR, and across several sensitivity analyses, increases confidence in these findings being causal.

A positive association between BMI and subfertility has been reported in observational studies among both women [3, 5, 8] and men [7, 8], with a particularly high risk of subfertility among obese individuals [4]. We confirm this association in our data, as higher BMI was associated with greater odds of subfertility from 22 kg/m^2^ in the standard multivariable regression models and from 23-25 kg/m^2^ in the MR analyses. These associations appeared unaffected by horizontal pleiotropy. Our findings are also supported by randomized controlled trials reporting an increase in overall pregnancies and natural conceptions among overweight/obese women after losing weight [9].

Several biological mechanisms can explain a potential association between high BMI and subfertility. Obesity is linked to biochemical disruptions (insulin resistance, adipocyte hyperactivation, greater levels of non-esterified fatty acids in plasma, increased hepatic triglyceride synthesis) [34]. These are in turn linked to impaired endocrine responses in women (lower synthesis of estrogens and luteinizing hormone, a greater production of androgens, and a decay in sex hormone binding globulins) and men (decreased testosterone levels, increased estrogen production in adipose tissue, defective hypothalamic pituitary gonadal regulation, decreased concentrations of sex hormone binding globulins) [34]. These endocrine alterations and other conditions linked to high BMI values, such as low-grade inflammation in reproductive tissues and some sex-dependent alterations (menstrual abnormalities, increased testicular heat, greater risk of erectile dysfunction), may finally compromise fecundity [2, 34-36].

The J-shaped association between BMI and subfertility also support that participants with low BMI may have a greater risk of subfertility. A decrease in BMI was linked to greater subfertility in women with a BMI <20 kg/m^2^, and we observed a similar tendency among men. Our results agree with previous observational studies reporting decreased fertility in underweight women who have undergone ART [6]. Low BMI values could be linked to subfertility because they are intimately related to undernutrition, which is associated with an impaired function of the reproductive system [37], defective concentrations of adipocyte-related regulators of endocrine processes such as leptin [38], and increased risk or pregnancy complications [39].

Our work presents some limitations. First, subfertility is a couple-dependent measure, and was reported by mothers in the cohort (if a woman was classified as subfertile, this condition was extrapolated to her partner). Thus, we are unable to determine whether subfertility was driven by the woman, man, or both. In addition, there is previous evidence of assortative mating on BMI [12], which could also confound the association between BMI and subfertility. Second, MoBa is a pregnancy cohort, and only includes couples who eventually conceived. Additional studies which are also able to include couples who never conceived are warranted. Third, the BMI GRS was associated with some predefined risk factors of subfertility, indicating that some horizontal pleiotropy may be present. However, multivariable MR and stratified analyses confirmed a robust association between BMI and subfertility, and additional sensitivity analyses found no evidence of horizontal pleiotropy in our data. Fourth, most of the associations with subfertility were found in the participants with extreme BMI values (of all women, 9.1% were obese and 12.4% had BMI values below 20.0 kg/m^2^; of all men, 10.2% were obese and only 1.25% had BMI values below 20.0 kg/m^2^) and therefore should be interpreted with caution. Finally, our study sample (couples who eventually conceived and were of a northern European ancestry) limits the generalizability of our conclusions to other populations. Nevertheless, our work also has several strengths. To our knowledge, studies exploring non-linear associations between BMI and subfertility using multivariable regressions and an MR approach have been lacking. Both present different sources of bias (multivariable regression could be biased by residual confounding, whilst MR could be biased by unbalanced horizontal pleiotropy), but the consistency in the findings according to both approaches increases confidence that these findings may be causal [14, 24]. This was facilitated by having large numbers of well-characterized participants with genome-wide and subfertility data coming from a relatively homogeneous population with northern European ancestry. This last aspect minimized the risk of confounding due to population stratification in our MR analyses, as well as the further adjustment for 10 ancestry-informative principal components [33]. Finally, our genetic instrument is robust [40, 41] and has been successfully used in several other MR studies [25, 42, 43].

## CONCLUSIONS

We observed a J-shaped relationship between BMI and subfertility in both sexes, when using both a standard multivariable regression and Mendelian randomization analysis. Taken together, our results support a causal role of BMI on subfertility. Current advice and support for overweight and obese subfertile couples to lose weight should continue. Clinicians should also consider appropriate advice to those with low BMI on how to increase their weight in a healthy way to promote fertility.

## Data Availability

The consent given by the participants does not open for storage of data on an individual level in repositories or journals. Researchers who want access to data sets for replication should submit an application to. Access to data sets requires approval from the Regional Committee for Medical and Health Research Ethics in Norway and an agreement with MoBa. Source data of the GWAS on BMI (Yengo L et al., Hum Mol Genet, 2018) are available in the GIANT Consortium website (https://portals.broadinstitute.org/collaboration/giant/index.php/GIANT_consortium_data_files#GWAS_Anthropometric_2015_BMI_Summary_Statistics). Source data of the GWAS on education years (Lee JJ et al., Nat Genet, 2018) are available in the Supplemental Tables of the article (https://www.nature.com/articles/s41588-018-0147-3#Sec34). Finally, source data of the GWAS on smoking initiation (Liu M et al., Nat Genet, 2019) are available in the Supplemental Tables of the article (https://www.nature.com/articles/s41588-018-0307-5#Sec14).

## ABBREVIATIONS

*ART*: assisted reproductive technology
*BMI*: body mass index
*GRS*: genetic risk score
*GWAS*: genome-wide association study
*MR*: Mendelian randomization
*MoBa*: the Norwegian Mother, Father, and Child cohort study
*OR*: odds ratio
*SNP*: single nucleotide polymorphism

## ACKNOWLEDGEMENTS

The MoBa Cohort Study is supported by the Norwegian Ministry of Health and Care Services and the Ministry of Education and Research. We are grateful to all the participating families in Norway who take part in this on-going cohort study, and those who contributed to the recruitment and the infrastructure surrounding the MoBa cohort.

We thank the Norwegian Institute of Public Health for generating high-quality genomic data. This research is part of the HARVEST collaboration, supported by the Research Council of Norway (#229624). We also thank the NORMENT Centre for providing genotype data, funded by the Research Council of Norway (#223273), South East Norway Health Authority and Stiftelsen Kristian Gerhard Jebsen. We further thank the Center for Diabetes Research (University of Bergen) for providing genotype information and performing quality control and imputation of the data in research projects funded by the European Research Council Advanced Grant SELECTionPREDISPOSED, Stiftelsen Kristian Gerhard Jebsen, the Trond Mohn Foundation, the Research Council of Norway, the Novo Nordisk Foundation, the University of Bergen, and the Western Norway Health Authority.

This work was performed on the TSD (Tjeneste for Sensitive Data) facilities, owned by the University of Oslo, operated and developed by the TSD service group at the University of Oslo, IT-Department.

This paper does not necessarily reflect the position or policy of the Norwegian Research Council.

## AUTHOR CONTRIBUTIONS

**Conceptualization:** Maria Christine Magnus

**Data curation:** Álvaro Hernáez, Maria Christine Magnus

**Formal analysis:** Álvaro Hernáez, Maria Christine Magnus

**Funding acquisition:** Maria Christine Magnus

**Investigation:** Siri E. Håberg, Abigail Fraser, Stephen Burgess, Deborah A. Lawlor, Maria Christine Magnus

**Methodology:** Álvaro Hernáez, Tormod Rogne, Karoline H. Skåra, Siri E. Håberg, Christian M. Page, Abigail Fraser, Stephen Burgess, Deborah A. Lawlor, Maria Christine Magnus

**Project administration:** Maria Christine Magnus

**Resources:** Maria Christine Magnus

**Software:** Álvaro Hernáez, Tormod Rogne, Karoline H. Skåra, Christian M. Page

**Supervision:** Siri E. Håberg, Abigail Fraser, Stephen Burgess, Deborah A. Lawlor, Maria Christine Magnus

**Validation:** Álvaro Hernáez

**Visualization:** Álvaro Hernáez, Karoline H. Skåra

**Writing – original draft:** Álvaro Hernáez

**Writing – review and editing:** Tormod Rogne, Karoline H. Skåra, Siri E. Håberg, Christian M. Page, Abigail Fraser, Stephen Burgess, Deborah A. Lawlor, Maria Christine Magnus

## FUNDING

The MoBa Cohort Study is supported by the Norwegian Ministry of Health and Care Services and the Norwegian Ministry of Education and Research. This project received funding from the European Research Council under the European Union’s Horizon 2020 research and innovation program (grant agreement No 947684). This work was also partly supported by the Research Council of Norway through its Centres of Excellence funding scheme, project number 262700. Open Access funding was provided by the Folkehelseinstituttet/Norwegian Institute of Public Health. D.A.L. is a UK National Institute for Health Research Senior Investigator (NF-SI-0611–10196) and is supported by the US National Institutes of Health (R01 DK10324) and a European Research Council Advanced Grant (DevelopObese; 669545). The funders had no role in the collection, analysis and interpretation of data; in the writing of the report; or in the decision to submit the article for publication.

## COMPETING INTERESTS

S.B. is a payed statistical reviewer for PLOS Medicine. D.A.L. receives (or has received in the last 10 years) research support from National and International government and charitable bodies, Roche Diagnostics and Medtronic for research unrelated to the current work. The rest of the authors declare that no competing interests exist.

## SUPPORTING INFORMATION

**S1 Checklist. STROBE-MR checklist for Mendelian randomization studies**.

**S2 Checklist. STROBE checklist for cohort studies**.

**S1 Fig. Association between body mass index and subfertility: multivariable non-linear Mendelian randomization considering the genetically determined number of education years in women (A) and men (B)**. A BMI of 25 kg/m^2^ was set as reference (black dot). Gray lines represent 95% confidence intervals.

**S2 Fig. Association between body mass index and subfertility: multivariable non-linear Mendelian randomization considering the genetic predisposition to having ever smoked in women (A) and men (B)**. A BMI of 25 kg/m^2^ was set as reference (black dot). Gray lines represent 95% confidence intervals.

**S3 Fig. Mendelian randomization analyses of body mass index and subfertility stratified by age**. Results in mothers below the median age (30 years, **A**) and over the median age (**B**), and of fathers below the median age (32 years, **C**) and over the median age (**D**) are presented. A BMI of 25 kg/m^2^ was set as reference (black dot). Gray lines represent 95% confidence intervals.

**S4 Fig. Scatterplot with the two sample Mendelian randomization analyses of body mass index and subfertility in women (A) and men (B)**. The figure also presents the MR estimates according to the inverse variance weighted regression, MR-Egger, weighted median method, and MR weighted mode estimator.

**S5 Fig. Mendelian randomization analyses of body mass index and subfertility in planned pregnancies in women (A) and men (B)**. A BMI of 25 kg/m^2^ was set as reference (black dot). Gray lines represent 95% confidence intervals.

**S6 Fig. Mendelian randomization analyses of body mass index and subfertility excluding assisted reproductive technology pregnancies in women (A) and men (B)**. A BMI of 25 kg/m^2^ was set as reference (black dot). Gray lines represent 95% confidence intervals.

**S1 Table. A priori subfertility risk factors tabulated against BMI genetic risk score quartiles in women**.

**S2 Table. A priori subfertility risk factors tabulated against BMI genetic risk score quartiles in men**.

**S3 Table. Two sample Mendelian randomization analyses of body mass index and subfertility, with indicators of horizontal pleiotropy and SNP heterogeneity**.

**S4 Table. Mendelian randomization analyses in planned pregnancies and after excluding assisted reproductive technology pregnancies**.

## Notes

### Author Declarations

The MoBa cohort is conducted according to the Declaration of Helsinki for Medical Research involving Human Subjects. The data collection in MoBa is approved by the Norwegian Data Inspectorate. Participants provided a written informed consent before joining the cohort. This project was approved by the Regional Committee for Medical and Health Research Ethics of South/East Norway (reference: 2017/1362).

## REFERENCES

1. Sallmén M, Sandler DP, Hoppin JA, Blair A, Baird DD. Reduced fertility among overweight and obese men. Epidemiology. 2006;17(5):520–3. doi: 10.1097/01.ede.0000229953.76862.e5.

2. Silvestris E, de Pergola G, Rosania R, Loverro G. Obesity as disruptor of the female fertility. Reprod Biol Endocrinol. 2018;16(1):22. doi: 10.1186/s12958-018-0336-z.

3. Ramlau-Hansen CH, Thulstrup AM, Nohr EA, Bonde JP, Sørensen TI, Olsen J. Subfecundity in overweight and obese couples. Hum Reprod. 2007;22(6):1634–7. doi: 10.1093/humrep/dem035.

4. van der Steeg JW, Steures P, Eijkemans MJ, Habbema JD, Hompes PG, Burggraaff JM, et al. Obesity affects spontaneous pregnancy chances in subfertile, ovulatory women. Hum Reprod. 2008;23(2):324–8. doi: 10.1093/humrep/dem371.

5. Pinborg A, Gaarslev C, Hougaard CO, Nyboe Andersen A, Andersen PK, Boivin J, et al. Influence of female bodyweight on IVF outcome: a longitudinal multicentre cohort study of 487 infertile couples. Reprod Biomed Online. 2011;23(4):490–9. doi: 10.1016/j.rbmo.2011.06.010.

6. Xiong YQ, Liu YM, Qi YN, Liu CR, Wang J, Li L, et al. Association between prepregnancy subnormal body weight and obstetrical outcomes after autologous in vitro fertilization cycles: systematic review and meta-analysis. Fertil Steril. 2020;113(2):344-53.e2. doi: 10.1016/j.fertnstert.2019.09.025.

7. Campbell JM, Lane M, Owens JA, Bakos HW. Paternal obesity negatively affects male fertility and assisted reproduction outcomes: a systematic review and meta-analysis. Reprod Biomed Online. 2015;31(5):593–604. doi: 10.1016/j.rbmo.2015.07.012.

8. Mushtaq R, Pundir J, Achilli C, Naji O, Khalaf Y, El-Toukhy T. Effect of male body mass index on assisted reproduction treatment outcome: an updated systematic review and meta-analysis. Reprod Biomed Online. 2018;36(4):459–71. doi: 10.1016/j.rbmo.2018.01.002.

9. Best D, Avenell A, Bhattacharya S. How effective are weight-loss interventions for improving fertility in women and men who are overweight or obese? A systematic review and meta-analysis of the evidence. Hum Reprod Update. 2017;23(6):681–705. doi: 10.1093/humupd/dmx027.

10. Collins GG, Rossi BV. The impact of lifestyle modifications, diet, and vitamin supplementation on natural fertility. Fertil Res Pract. 2015;1:11. doi: 10.1186/s40738-015-0003-4.

11. Hart RJ. Physiological Aspects of Female Fertility: Role of the Environment, Modern Lifestyle, and Genetics. Physiol Rev. 2016;96(3):873–909. doi: 10.1152/physrev.00023.2015.

12. Silventoinen K, Kaprio J, Lahelma E, Viken RJ, Rose RJ. Assortative mating by body height and BMI: Finnish twins and their spouses. Am J Hum Biol. 2003;15(5):620–7. doi: 10.1002/ajhb.10183.

13. Lawlor DA, Harbord RM, Sterne JA, Timpson N, Davey Smith G. Mendelian randomization: using genes as instruments for making causal inferences in epidemiology. Stat Med. 2008;27(8):1133–63. doi: 10.1002/sim.3034.

14. Davey Smith G, Hemani G. Mendelian randomization: genetic anchors for causal inference in epidemiological studies. Hum Mol Genet. 2014;23(R1):R89–98. doi: 10.1093/hmg/ddu328.

15. Lawlor DA, Tilling K, Davey Smith G. Triangulation in aetiological epidemiology. Int J Epidemiol. 2016;45(6):1866–86. doi: 10.1093/ije/dyw314.

16. Magnus P, Birke C, Vejrup K, Haugan A, Alsaker E, Daltveit AK, et al. Cohort Profile Update: The Norwegian Mother and Child Cohort Study (MoBa). Int J Epidemiol. 2016;45(2):382–8. doi: 10.1093/ije/dyw029.

17. Magnus P, Irgens LM, Haug K, Nystad W, Skjaerven R, Stoltenberg C. Cohort profile: the Norwegian Mother and Child Cohort Study (MoBa). Int J Epidemiol. 2006;35(5):1146–50. doi: 10.1093/ije/dyl170.

18. Paltiel L, Haugan A, Skjerden T K; H, Bækken S, Stensrud NK, et al. The biobank of the Norwegian Mother and Child Cohort Study – present status. Norsk Epidemiologi. 2014;24(1-2):29–35. doi: 10.5324/nje.v24i1-2.1755.

19. Helgeland Ø, Vaudel M, Juliusson PB, Lingaas Holmen O, Juodakis J, Bacelis J, et al. Genome-wide association study reveals dynamic role of genetic variation in infant and early childhood growth. Nat Commun. 2019;10(1):4448. doi: 10.1038/s41467-019-12308-0.

20. Yengo L, Sidorenko J, Kemper KE, Zheng Z, Wood AR, Weedon MN, et al. Meta-analysis of genome-wide association studies for height and body mass index in ∼700000 individuals of European ancestry. Hum Mol Genet. 2018;27(20):3641–9. doi: 10.1093/hmg/ddy271.

21. Choi SW, Mak TS, O’Reilly PF. Tutorial: a guide to performing polygenic risk score analyses. Nat Protoc. 2020;15(9):2759–72. doi: 10.1038/s41596-020-0353-1.

22. Barrabés NØ, Greta Kjølstad. Norsk standard for utdanningsgruppering. 2016 [Accessed March 17, 2021]. Available from: https://www.ssb.no/utdanning/_attachment/283616?_ts=1583e453200.

23. Rietveld CA, Medland SE, Derringer J, Yang J, Esko T, Martin NW, et al. GWAS of 126,559 individuals identifies genetic variants associated with educational attainment. Science. 2013;340(6139):1467–71. doi: 10.1126/science.1235488.

24. Sun YQ, Burgess S, Staley JR, Wood AM, Bell S, Kaptoge SK, et al. Body mass index and all cause mortality in HUNT and UK Biobank studies: linear and non-linear mendelian randomisation analyses. BMJ. 2019;364:1042. doi: 10.1136/bmj.l1042.

25. Rogne T, Solligård E, Burgess S, Brumpton BM, Paulsen J, Prescott HC, et al. Body mass index and risk of dying from a bloodstream infection: A Mendelian randomization study. PLoS Med. 2020;17(11):e1003413. doi: 10.1371/journal.pmed.1003413.

26. Burgess S, Davey Smith G, Davies NM, Dudbridge F, Gill D, Glymour MM, et al. Guidelines for performing Mendelian randomization investigations. Wellcome Open Res. 2019;4:186. doi: 10.12688/wellcomeopenres.15555.2.

27. Burgess S, Thompson SG. Multivariable Mendelian randomization: the use of pleiotropic genetic variants to estimate causal effects. Am J Epidemiol. 2015;181(4):251–60. doi: 10.1093/aje/kwu283.

28. Lee JJ, Wedow R, Okbay A, Kong E, Maghzian O, Zacher M, et al. Gene discovery and polygenic prediction from a genome-wide association study of educational attainment in 1.1 million individuals. Nat Genet. 2018;50(8):1112–21. doi: 10.1038/s41588-018-0147-3.

29. Liu M, Jiang Y, Wedow R, Li Y, Brazel DM, Chen F, et al. Association studies of up to 1.2 million individuals yield new insights into the genetic etiology of tobacco and alcohol use. Nat Genet. 2019;51(2):237–44. doi: 10.1038/s41588-018-0307-5.

30. Bowden J, Davey Smith G, Burgess S. Mendelian randomization with invalid instruments: effect estimation and bias detection through Egger regression. Int J Epidemiol. 2015;44(2):512–25. doi: 10.1093/ije/dyv080.

31. Bowden J, Davey Smith G, Haycock PC, Burgess S. Consistent Estimation in Mendelian Randomization with Some Invalid Instruments Using a Weighted Median Estimator. Genet Epidemiol. 2016;40(4):304–14. doi: 10.1002/gepi.21965.

32. Hemani G, Bowden J, Davey Smith G. Evaluating the potential role of pleiotropy in Mendelian randomization studies. Hum Mol Genet. 2018;27(R2):R195–r208. doi: 10.1093/hmg/ddy163.

33. Wang C, Zhan X, Liang L, Abecasis GR, Lin X. Improved ancestry estimation for both genotyping and sequencing data using projection procrustes analysis and genotype imputation. Am J Hum Genet. 2015;96(6):926–37. doi: 10.1016/j.ajhg.2015.04.018.

34. Amiri M, Ramezani Tehrani F. Potential Adverse Effects of Female and Male Obesity on Fertility: A Narrative Review. Int J Endocrinol Metab. 2020;18(3):e101776. doi: 10.5812/ijem.101776.

35. Broughton DE, Moley KH. Obesity and female infertility: potential mediators of obesity’s impact. Fertil Steril. 2017;107(4):840–7. doi: 10.1016/j.fertnstert.2017.01.017.

36. Salas-Huetos A, Maghsoumi-Norouzabad L, James ER, Carrell DT, Aston KI, Jenkins TG, et al. Male adiposity, sperm parameters and reproductive hormones: An updated systematic review and collaborative meta-analysis. Obes Rev. 2021;22(1):e13082. doi: 10.1111/obr.13082.

37. Cai J, Liu L, Zhang J, Qiu H, Jiang X, Li P, et al. Low body mass index compromises live birth rate in fresh transfer in vitro fertilization cycles: a retrospective study in a Chinese population. Fertil Steril. 2017;107(2):422-9.e2. doi: 10.1016/j.fertnstert.2016.10.029.

38. Mitchell M, Armstrong DT, Robker RL, Norman RJ. Adipokines: implications for female fertility and obesity. Reproduction. 2005;130(5):583–97. doi: 10.1530/rep.1.00521.

39. Dickey RP, Xiong X, Xie Y, Gee RE, Pridjian G. Effect of maternal height and weight on risk for preterm singleton and twin births resulting from IVF in the United States, 2008-2010. Am J Obstet Gynecol. 2013;209(4):349.e1-6. doi: 10.1016/j.ajog.2013.05.052.

40. Burgess S, Thompson SG. Bias in causal estimates from Mendelian randomization studies with weak instruments. Stat Med. 2011;30(11):1312–23. doi: 10.1002/sim.4197.

41. Evans DM, Brion MJ, Paternoster L, Kemp JP, McMahon G, Munafò M, et al. Mining the human phenome using allelic scores that index biological intermediates. PLoS Genet. 2013;9(10):e1003919. doi: 10.1371/journal.pgen.1003919.

42. Cheung CL, Tan KCB, Au PCM, Li GHY, Cheung BMY. Evaluation of GDF15 as a therapeutic target of cardiometabolic diseases in human: A Mendelian randomization study. EBioMedicine. 2019;41:85–90. doi: 10.1016/j.ebiom.2019.02.021.

43. Takahashi H, Cornish AJ, Sud A, Law PJ, Disney-Hogg L, Calvocoressi L, et al. Mendelian randomization provides support for obesity as a risk factor for meningioma. Sci Rep. 2019;9(1):309. doi: 10.1038/s41598-018-36186-6.

