## Supplemental Materials for "Body mass index and subfertility: multivariable regression and Mendelian randomization analyses in the Norwegian Mother, Father and Child Cohort Study"

**CHECKLISTS**

**S1 Checklist. STROBE-MR checklist for Mendelian randomization studies**

| Item | Complete/location |
|---|---|
| **Title and Abstract** | |
| **1. Title and Abstract:** "Mendelian randomization" is named both in the title and the abstract | Complete |
| **Introduction** | |
| **2. Background:** Explain the scientific background and rationale for the reported study. Is causality between exposure and outcome plausible? Justify why MR is a helpful method to address the study question. | Introduction, paragraph #1 |
| **3. Objectives:** State specific objectives clearly, including pre-specified causal hypotheses (if any). | Introduction, paragraph #2 |
| **Methods** | |
| **4. Study design and data sources:** Present key elements of study design early in the paper. Consider including a table listing sources of data for all phases of the study. For each data source contributing to the analysis, describe the following:<br>a) Describe the study design and the underlying population from which it was drawn. Describe also the setting, locations, and relevant dates, including periods of recruitment, exposure, follow-up, and data collection, if available.<br>b) Give the eligibility criteria, and the sources and methods of selection of participants.<br>c) Explain how the analyzed sample size was arrived at.<br>d) Describe measurement, quality and selection of genetic variants.<br>e) For each exposure, outcome and other relevant variables, describe methods of assessment and, in the case of diseases, the diagnostic criteria used.<br>f) Provide details of ethics committee approval and participant informed consent, if relevant. | a) Methods, "*The Norwegian Mother, Father and Child Cohort Study"* subsection<br>b) Methods, "*The Norwegian Mother, Father and Child Cohort Study"* subsection<br>c) Figure 1<br>d) Methods, "*Polygenic risk score BMI"* subsection<br>e) Methods, "*BMI*", "*Polygenic risk score for BMI, "Subfertility",* and *"Other variables"* subsections<br>f) Methods, "*Ethical approval*" subsection |
| **5. Assumptions:** Explicitly state assumptions for the main analysis (e.g. relevance, exclusion, independence, homogeneity) as well assumptions for any additional or sensitivity analysis. | Methods, "*Statistical analyses*" subsection, paragraph #2 |
| **6. Statistical methods**<br>Describe statistical methods and statistics used.<br>a) Describe how quantitative variables were handled in the analyses (i.e., scale, units, model). | a) Methods, "*Statistical analyses*" subsection, paragraph #1<br>b) Methods, "*Polygenic risk score for BMI"* subsection and "*Statistical analyses*" subsection, paragraph #3 |

| | |
|---|---|
| b) Describe the process for identifying genetic variants and weights to be included in the analyses (i.e, independence and model). Consider a flow diagram.<br>c) Describe the MR estimator, e.g. two-stage least squares, Wald ratio, and related statistics. Detail the included covariates and, in case of two-sample MR, whether the same covariate set was used for adjustment in the two samples.<br>d) Explain how missing data were addressed.<br>e) If applicable, say how multiple testing was dealt with. | c) Methods, "*Statistical analyses*" subsection, paragraph #3<br>d) Methods, "*Statistical analyses*" subsection, paragraph #6<br>e) Not applicable |
| **7. Assessment of assumptions**<br>Describe any methods used to assess the assumptions or justify their validity. | Methods, "*Statistical analyses*" subsection, paragraphs #3, #4, #5, and #6 |
| **8. Sensitivity analyses:** Describe any sensitivity analyses or additional analyses performed. | Methods, "*Statistical analyses*" subsection, paragraphs #5, #6, and #7 |
| **9. Software and pre-registration**<br>a) Name statistical software and package(s).<br>b) State whether the study protocol and details were pre-registered (as well as when and where). | Methods, "*Statistical analyses*" subsection, paragraph #8 |
| **Results** | |
| **10. Descriptive data**<br>a) Report the numbers of individuals at each stage of included studies and reasons for exclusion. Consider use of a flow-diagram.<br>b) Report summary statistics for phenotypic exposure(s), outcome(s) and other relevant variables (e.g. means, standard deviations, proportions).<br>c) If the data sources include meta-analyses of previous studies, provide the number of studies, their reported ancestry, if available, and assessments of heterogeneity across these studies. Consider using a supplementary table for each data source.<br>d) For two-sample Mendelian randomization:<br>i. Provide information on the similarity of the genetic variant-exposure associations between the exposure and outcome samples.<br>ii. Provide information on extent of sample overlap between the exposure and outcome data sources. | a) Figure 1; Results, "*Study population*" subsection<br>b) Table 1; Results, "*Study population*" and "*Association between reported BMI and subfertility*" subsections<br>c) Not applicable<br>d) Methods, "*Statistical analyses*" subsection, paragraph #6 |
| **11. Main results**<br>a) Report the associations between genetic variant and exposure, and between genetic variant and outcome, preferably on an interpretable scale<br>b) Report causal effect estimate between exposure and outcome, and the measures of uncertainty from the MR | a) Genetic variants-exposure: Results, "*Strength of the genetic instrument for BMI", "MR analyses on BMI and subfertility in women",* and *"MR analyses on BMI and subfertility in men"* subsections. |

| | |
|---|---|
| analysis. Use an intuitive scale, such as odds ratio, or relative risk, per standard deviation difference.<br>c) If relevant, consider translating estimates of relative risk into absolute risk for a meaningful time-period.<br>d) Consider any plots to visualize results (e.g. forest plot, scatterplot of associations between genetic variants and outcome versus between genetic variants and exposure). | Genetic variants-outcome: Results, *"MR analyses on BMI and subfertility in women",* and *"MR analyses on BMI and subfertility in men"* subsections.<br>b) ORs [95% CI] are used<br>c) Not applicable<br>d) Figures 2, 3 and 4 |
| **12. Assessment of assumptions**<br>a) Assess the validity of the assumptions.<br>b) Report any additional statistics (e.g., assessments of heterogeneity, such as I2, Q statistic). | Results, "*Evaluation of horizontal pleiotropy*". |
| **13. Sensitivity and additional analyses**<br>a) Use sensitivity analyses to assess the robustness of the main results to violations of the assumptions.<br>b) Report results from other sensitivity analyses (e.g., replication study with different dataset, analyses of subgroups, validation of instrument(s), simulations, etc.).<br>c) Report any assessment of direction of causality (e.g., bidirectional MR).<br>d) When relevant, report and compare with estimates from non-MR analyses.<br>e) Consider any additional plots to visualize results (e.g., leave-one-out analyses). | a) Results, "*Evaluation of horizontal pleiotropy*"; Table 2; Supplemental figures S1-S4; Supplemental Tables S1 and S2.<br>b) Results, *"Other sensitivity analyses"* subsections; Table 2; Supplemental figures S1-S6; Supplemental Tables S1 and S2<br>c) Not applicable<br>d) Results, "*Association between reported BMI and subfertility"*<br>e) Supplemental Figures S1-S6 |
| **Discussion** | |
| **14. Key results** | Discussion, paragraph #1; Conclusions. |
| **15. Limitations**<br>Discuss limitations of the study, taking into account the validity of the MR assumptions, other sources of potential bias, and imprecision. Discuss both direction and magnitude of any potential bias, and any efforts to address them. | Discussion, paragraph #4 |
| **16. Interpretations**<br>a) Give a cautious overall interpretation of results considering objectives and limitations. Compare with results from other relevant studies.<br>b) Discuss underlying biological mechanisms that could be modelled by using the genetic<br>variants to assess the relationship between the exposure and the outcome.<br>c) Discuss whether the results have clinical or policy relevance, and whether interventions could have the same size effect. | a) Discussion, paragraphs #2 and #3<br>b) Discussion, paragraphs #2 and #3<br>c) Discussion, paragraphs #2 and #3; Conclusions |
| **17. Generalizability** | Discussion, paragraph #4; Conclusions. |
| **18. Funding** | Funding (page 26) |
| **19. Data and data sharing** | Data availability (page 25) |
| **20. Conflicts of Interest** | Competing interests (page 26) |

**S2 Checklist. STROBE checklist for cohort studies**

| | Item No | Recommendation | Page No |
|---|---|---|---|
| **Title and abstract** | | | |
| Title and abstract | 1 | (*a*) Indicate the study's design with a commonly used term in the title or the abstract | 1 |
| | | (*b*) Provide in the abstract an informative and balanced summary of what was done and what was found | 2-3 |
| **Introduction** | | | |
| Background/ rationale | 2 | Explain the scientific background and rationale for the investigation being reported | 6 |
| Objectives | 3 | State specific objectives, including any pre-specified hypotheses | 6 |
| **Methods** | | | |
| Study design | 4 | Present key elements of study design early in the paper | 6-7 |
| Setting | 5 | Describe the setting, locations, and relevant dates, including periods of recruitment, exposure, follow-up, and data collection | 6-7 |
| Participants | 6 | (*a*) Give the eligibility criteria, and the sources and methods of selection of participants. Describe methods of follow-up | 6-7 |
| | | (*b*) For matched studies, give matching criteria and number of exposed and unexposed | - |
| Variables | 7 | Clearly define all outcomes, exposures, predictors, potential confounders, and effect modifiers. Give diagnostic criteria, if applicable | 7-8 |
| Data sources/ measurement | 8 | For each variable of interest, give sources of data and details of methods of assessment (measurement). Describe comparability of assessment methods if there is more than one group | 7-8 |
| Bias | 9 | Describe any efforts to address potential sources of bias | 8-12 |
| Study size | 10 | Explain how the study size was arrived at | Fig.1 |
| Quantitative variables | 11 | Explain how quantitative variables were handled in the analyses. If applicable, describe which groupings were chosen and why | 7-8 |
| Statistical methods | 12 | (*a*) Describe all statistical methods, including those used to control for confounding | 9-12 |
| | | (*b*) Describe any methods used to examine subgroups and interactions | 9-12 |
| | | (*c*) Explain how missing data were addressed | - |
| | | (*d*) If applicable, explain how loss to follow-up was addressed | - |
| | | (*e*) Describe any sensitivity analyses | - |
| **Results** | | | |
| Participants | 13* | (*a*) Report numbers of individuals at each stage of study—e.g. numbers potentially eligible, examined for eligibility, confirmed eligible, included in the study, completing follow-up, and analyzed | 13-14, Fig.1 |
| | | (*b*) Give reasons for non-participation at each stage | |
| | | (*c*) Consider use of a flow diagram | |

| | | | |
|---|---|---|---|
| Descriptive data | 14* | (*a*) Give characteristics of study participants (e.g. demographic, clinical, social) and information on exposures and potential confounders | 13-14, Table 1 |
| | | (*b*) Indicate number of participants with missing data for each variable of interest | - |
| | | (*c*) Summarize follow-up time (e.g., average and total amount) | 6-7 |
| Outcome data | 15 | Report numbers of outcome events or summary measures over time | 13 |
| Main results | 16 | (*a*) Give unadjusted estimates and, if applicable, confounder-adjusted estimates and their precision (e.g., 95% confidence interval). Make clear which confounders were adjusted for and why they were included | 14-21 |
| | | (*b*) Report category boundaries when continuous variables were categorized | 14-21 |
| | | (*c*) If relevant, consider translating estimates of relative risk into absolute risk for a meaningful time period | - |
| Other analyses | 17 | Report other analyses done—e.g. analyses of subgroups and interactions, and sensitivity analyses | 14-21 |
| **Discussion** | | | |
| Key results | 18 | Summarize key results with reference to study objectives | 21, 24 |
| Limitations | 19 | Discuss limitations of the study, taking into account sources of potential bias or imprecision. Discuss both direction and magnitude of any potential bias | 22-23 |
| Interpretation | 20 | Give a cautious overall interpretation of results considering objectives, limitations, multiplicity of analyses, results from similar studies, and other relevant evidence | 22-23 |
| Generalizability | 21 | Discuss the generalizability (external validity) of the study results | 23 |
| **Other information** | | | |
| Funding | 22 | Give the source of funding and the role of the funders for the present study and, if applicable, for the original study on which the present article is based | 26 |

7

### SUPPORTING INFORMATION

### SUPPLEMENTAL METHODS

*GWAS on subfertility in the MoBa cohort*

The instrumental variables linked to subfertility were defined in a GWAS performed in the MoBa cohort. We analyzed the association between the genotyped SNPs in MoBa after the quality control process [1] and subfertility using logistic regressions in women and men separately. Considering that some parents were involved in the cohort with more than one pregnancy, any participant reporting a time-to-pregnancy ≥12 months or the use of assisted reproductive technologies in any of their pregnancies was considered subfertile. The MoBa Genetics infrastructure is a collaborative research effort organized in five genotype batches of parent-offspring trios [1, 2]. Thus, we assessed the association between SNPs and subfertility in each batch using Plink [3] and finally performed a sample size-weighted meta-analysis using a random effects model by the GWAMA software [4]. No MoBa participant was involved in the GWAS that defined the genetic instrumental variables for the exposure (both populations were completely independent).

### SUPPLEMENTAL RESULTS

*Validity of genetic instruments for education years and smoking initiation*

The genetic instrument of education years was robust. Each one unit increase in the GRS was linked to an increase of 0.028 education years in women (95% CI 0.027 to 0.030, $p < 0.001$, 4.03% of education year variation explained, *F*-statistic = 1,082) and 0.032 education years in men (95% CI 0.030 to 0.034, $p < 0.001$, 4.27% of education year variation explained, *F*-statistic = 1,070).

The genetic instrument of the likelihood of having ever smoked was also robust. Each one unit increase in the GRS was associated with 2% greater odds of having ever initiated smoking in women (OR 1.02, 95% CI 1.019 to 1.022, $p < 0.001$, 1.89% of variance explained, area under the ROC curve = 0.567) and men (OR 1.02, 95% CI 1.018 to 1.022, $p < 0.001$, 1.52% of variance explained, area under the ROC curve = 0.553).

**SUPPLEMENTAL FIGURES**

**S1 Fig. Association between body mass index and subfertility: multivariable non-linear Mendelian randomization considering the genetically determined number of education years in women (A) and men (B).** A BMI of 25 kg/$m^2$ was set as reference (black dot). Gray lines represent 95% confidence intervals.

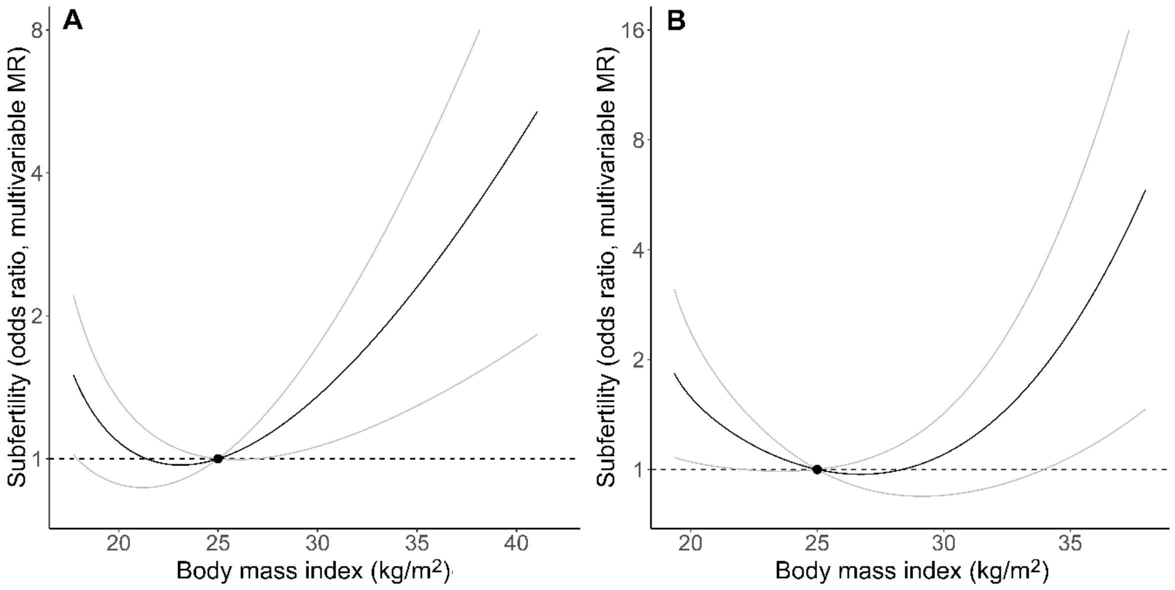

**S2 Fig. Association between body mass index and subfertility: multivariable non-linear Mendelian randomization considering the genetic predisposition to having ever smoked in women (A) and men (B).** A BMI of 25 kg/m$^2$ was set as reference (black dot). Gray lines represent 95% confidence intervals.

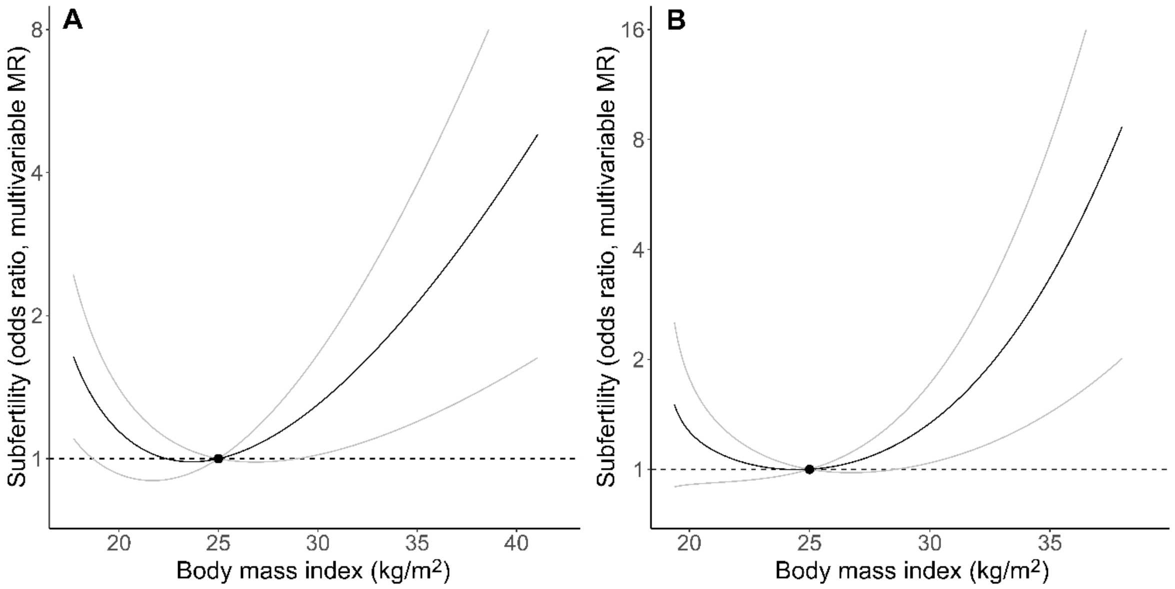

**S3 Fig. Mendelian randomization analyses of body mass index and subfertility stratified by age.** Results in mothers below the median age (30 years, **A**) and over the median age (**B**), and of fathers below the median age (32 years, **C**) and over the median age (**D**) are presented. A BMI of 25 kg/m$^2$ was set as reference (black dot). Gray lines represent 95% confidence intervals.

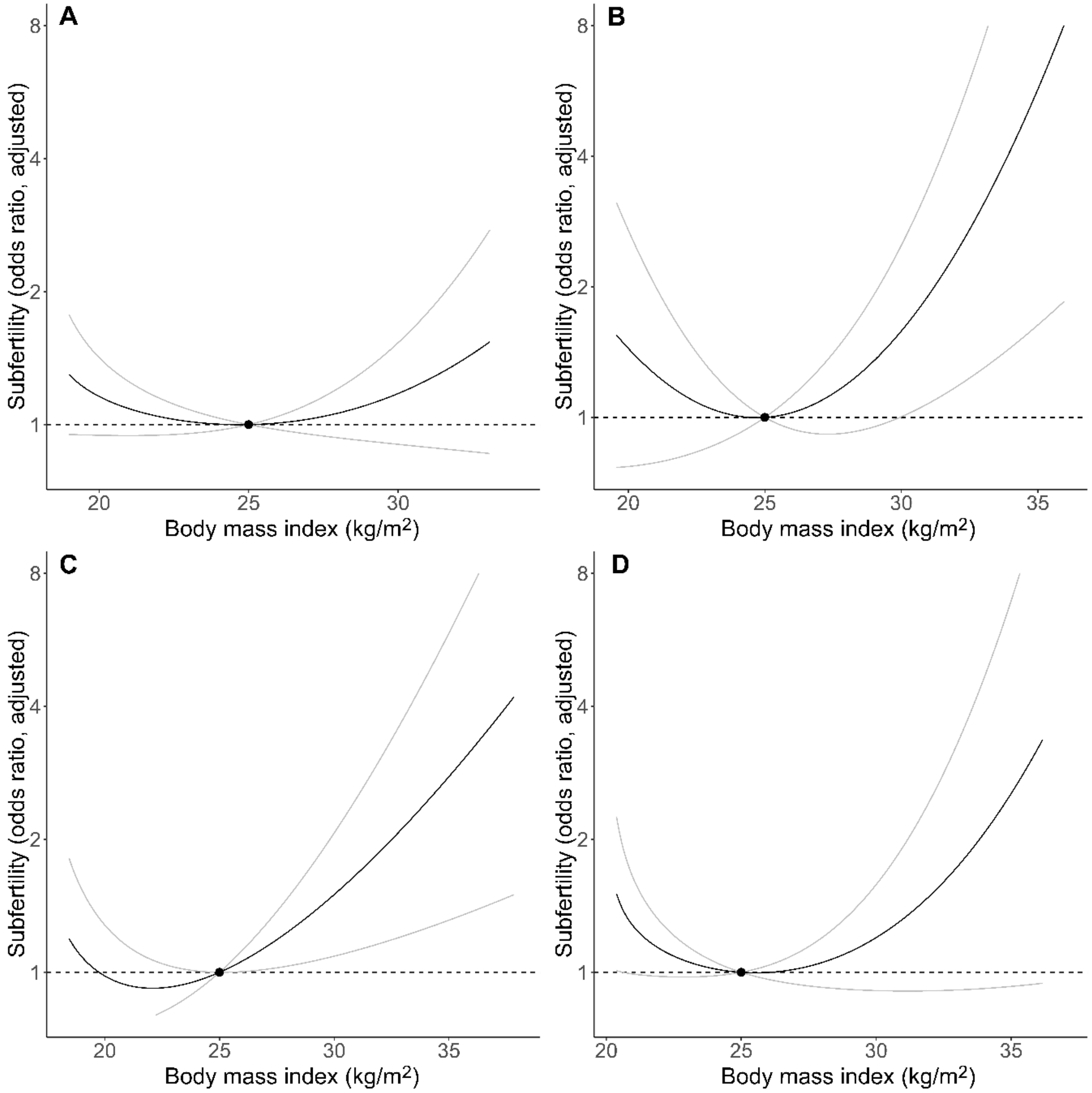

**S4 Fig. Scatterplot with the two sample Mendelian randomization analyses of body mass index and subfertility in women (A) and men (B).** The figure also presents the MR estimates according to the inverse variance weighted regression, MR-Egger, weighted median method, and MR weighted mode estimator.

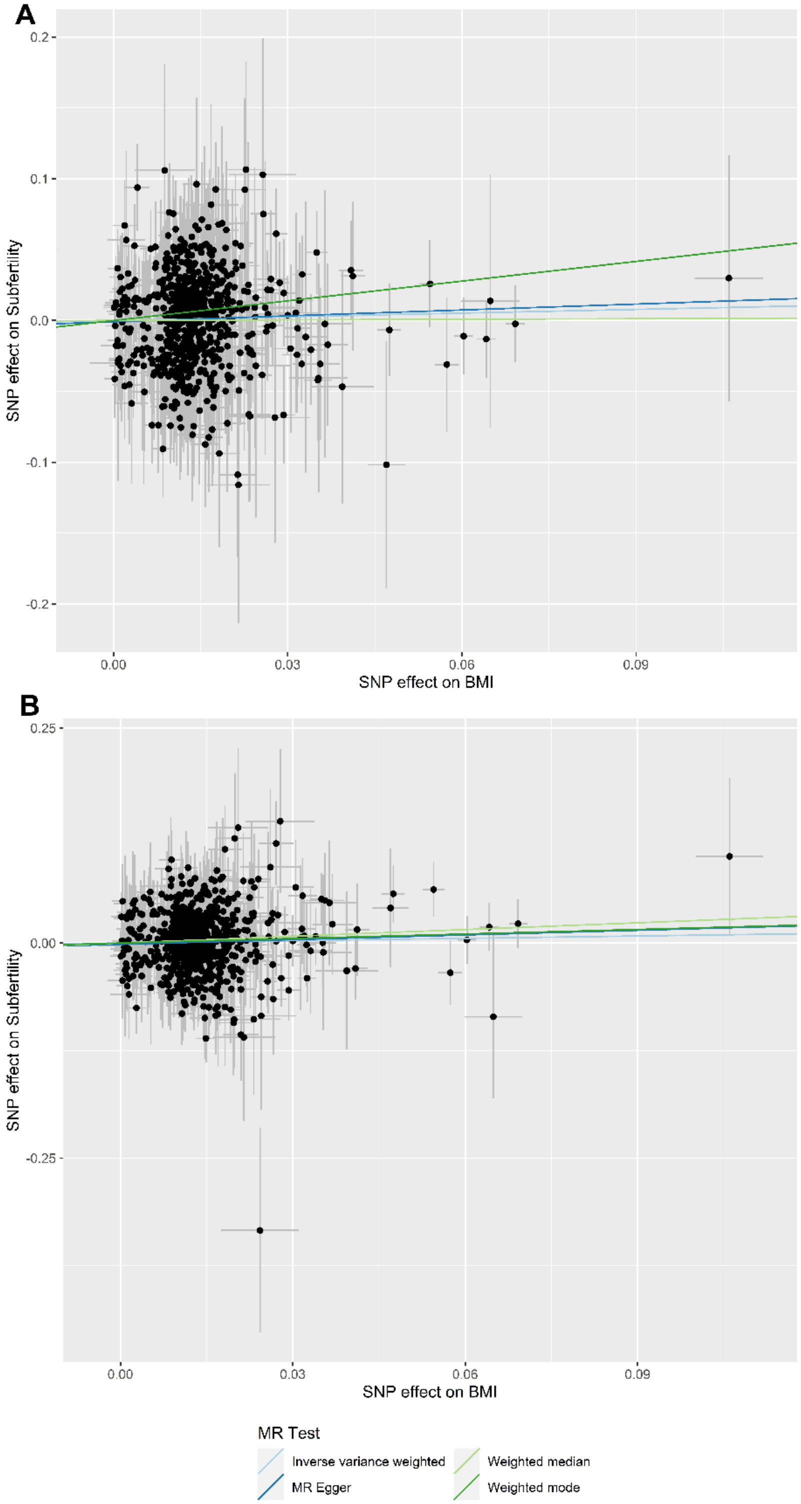

**S5 Fig. Mendelian randomization analyses of body mass index and subfertility in planned pregnancies in women (A) and men (B).** A BMI of 25 kg/m$^2$ was set as reference (black dot). Gray lines represent 95% confidence intervals.

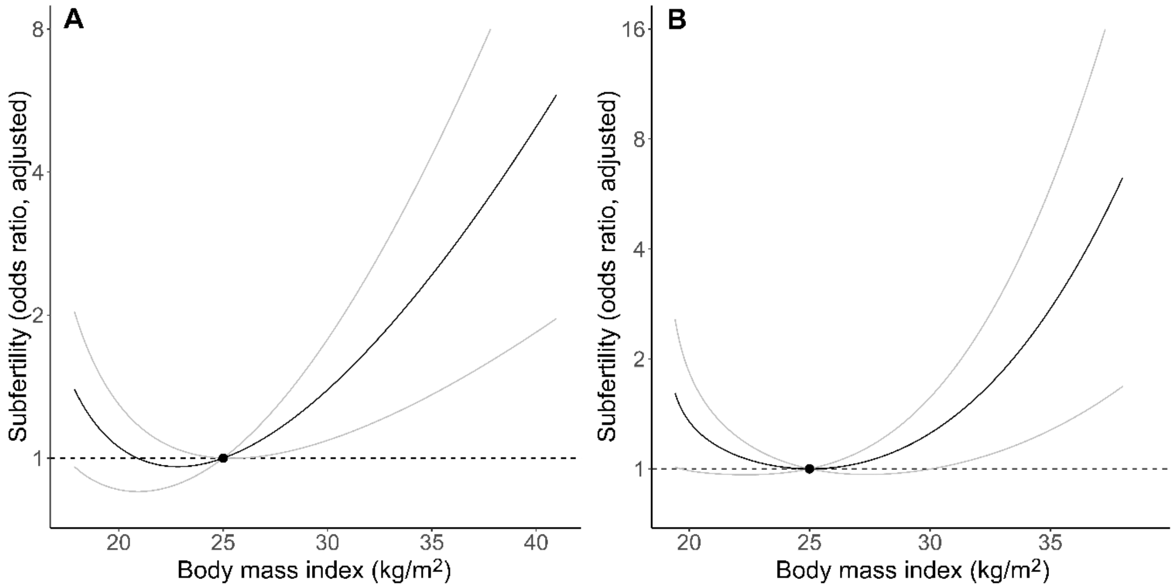

**S6 Fig. Mendelian randomization analyses of body mass index and subfertility excluding assisted reproductive technology pregnancies in women (A) and men (B).** A BMI of 25 kg/m$^2$ was set as reference (black dot). Gray lines represent 95% confidence intervals.

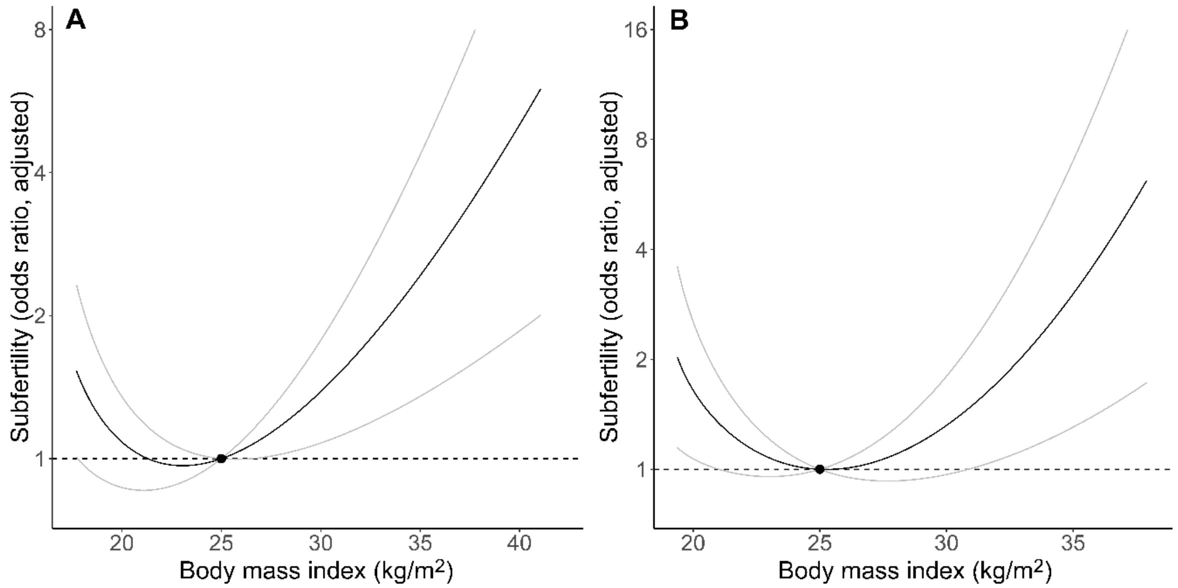

### SUPPLEMENTAL TABLES

**S1 Table. A priori subfertility risk factors tabulated against BMI genetic risk score quartiles in women.**

| | **Quartiles according to BMI genetic risk scores** | | | | *p*-value (linear trend) | *p*-value (chi-squared) |
|---|---|---|---|---|---|---|
| | Quartile 1 (*n* = 8528) | Quartile 2 (*n* = 8529) | Quartile 3 (*n* = 8525) | Quartile 4 (*n* = 8523) | | |
| Age, years (mean ± SD) | 30.2 ± 4.32 | 30.1 ± 4.38 | 30.1 ± 4.40 | 30.0 ± 4.39 | <0.001 | - |
| Education years (mean ± SD) | 17.5 ± 3.16 | 17.4 ± 3.17 | 17.3 ± 3.21 | 17.3 ± 3.23 | <0.001 | - |
| Tobacco use (*n*, %): | | | | | - | <0.001 |
| Never smokers | 4563 (53.6%) | 4620 (54.4%) | 4361 (51.2%) | 4340 (51.0%) | | |
| Current/former smokers | 3948 (46.4%) | 3876 (45.6%) | 4150 (48.8%) | 4165 (49.0%) | | |
| Previous pregnancies (*n*, %): | | | | | - | 0.200 |
| 0 | 3905 (45.8%) | 3969 (46.5%) | 3842 (45.1%) | 3852 (45.2%) | | |
| ≥1 | 4623 (54.2%) | 4560 (53.5%) | 4683 (54.9%) | 4671 (54.8%) | | |

SD: standard deviation.

**S2 Table. A priori subfertility risk factors tabulated against BMI genetic risk score quartiles in men**

| | **Quartiles according to BMI genetic risk scores** | | | | *p*-value (linear trend) | *p*-value (chi-squared) |
|---|---|---|---|---|---|---|
| | Quartile 1 (*n* = 7869) | Quartile 2 (*n* = 7859) | Quartile 3 (*n* = 7861) | Quartile 4 (*n* = 7866) | | |
| Age, years (mean ± SD) | 32.8 ± 5.20 | 32.6 ± 5.00 | 32.5 ± 5.12 | 32.4 ± 5.11 | <0.001 | - |
| Education years (mean ± SD) | 16.6 ± 3.51 | 16.5 ± 3.53 | 16.4 ± 3.54 | 16.3 ± 3.53 | <0.001 | - |
| Tobacco use (*n*, %): | | | | | - | <0.001 |
| Never smokers | 5906 (75.2%) | 5790 (73.8%) | 5674 (72.3%) | 5628 (71.7%) | | |
| Current/former smokers | 1945 (24.8%) | 2053 (26.2%) | 2170 (27.7%) | 2219 (28.3%) | | |
| Previous pregnancies (*n*, %): | | | | | - | 0.531 |
| 0 | 3583 (45.5%) | 3589 (45.7%) | 3633 (46.2%) | 3663 (46.6%) | | |
| ≥1 | 4286 (54.5%) | 4270 (54.3%) | 4228 (53.8%) | 4203 (53.4%) | | |

SD: standard deviation.

**S3 Table. Two sample Mendelian randomization analyses of body mass index and subfertility, with indicators of horizontal pleiotropy and SNP heterogeneity.**

| MR method | Beta (standard error) | *p*-value | Heterogeneity (*Q*, *Q'*) | Heterogeneity (*p*-value) | Horizonal pleiotropy (*p*-value) |
|---|---|---|---|---|---|
| **Women** | | | | | |
| Inverse variance weighted | -0.11 (0.075) | 0.137 | 719.85 | 0.999 | - |
| MR Egger | -0.15 (0.15) | 0.337 | 719.77 | 0.999 | 0.780 |
| Weighted median | -0.085 (0.13) | 0.509 | - | - | - |
| Weighted mode | -0.019 (1.73) | 0.991 | - | - | - |
| **Men** | | | | | |
| Inverse variance weighted | -0.052 (0.077) | 0.496 | 790.63 | 0.968 | - |
| MR Egger | -0.14 (0.16) | 0.369 | 790.21 | 0.967 | 0.514 |
| Weighted median | -0.23 (0.15) | 0.113 | - | - | - |
| Weighted mode | -0.18 (2.12) | 0.934 | - | - | - |

*MR*: Mendelian randomization; *Q*: Cochran's *Q* statistic (heterogeneity according to the inverse variance weighted regression); *Q'*: Rücker's *Q'* statistic (heterogeneity according to the MR-Egger method)

**S4 Table. Mendelian randomization analyses in planned pregnancies and after excluding assisted reproductive technology pregnancies.**

| | MR: main analyses | MR: only participants in planned pregnancies | MR: only non-ART conceptions in subfertile cases |
|---|---|---|---|
| **Women** | | | |
| ***Linear MR*** | | | |
| OR for $\Delta$1 kg/m$^2$ (whole population) | 1.02 (0.99 to 1.06) | 1.04 (1.00 to 1.08) | 1.04 (1.00 to 1.08) |
| ***Non-linear MR*** | | | |
| Fractional polynomial test (*p*-value) | 0.033 | 0.030 | 0.028 |
| OR for $\Delta$1 kg/m$^2$ (stratified analyses): | | | |
| < 20.0 kg/m$^2$ | 0.86 (0.75 to 0.97) | 0.86 (0.76 to 0.98) | 0.88 (0.77 to 1.02) |
| 20.0-24.9 kg/m$^2$ | 0.99 (0.94 to 1.04) | 1.01 (0.96 to 1.06) | 0.98 (0.93 to 1.04) |
| 25.0-29.9 kg/m$^2$ | 1.03 (0.96 to 1.11) | 1.05 (0.97 to 1.13) | 1.05 (0.97 to 1.14) |
| ≥ 30.0 kg/m$^2$ | 1.15 (1.04 to 1.28) | 1.17 (1.05 to 1.31) | 1.20 (1.07 to 1.34) |
| BMI with lowest subfertility odds | 23.3 kg/m$^2$ | 22.8 kg/m$^2$ | 23.1 kg/m$^2$ |
| **Men** | | | |
| ***Linear MR*** | | | |
| OR for $\Delta$1 kg/m$^2$ (whole population) | 1.01 (0.96 to 1.07) | 1.02 (0.97 to 1.08) | 1.03 (0.97 to 1.09) |
| ***Non-linear MR*** | | | |
| Fractional polynomial test (*p*-value) | 0.018 | 0.014 | 0.022 |
| OR for $\Delta$1 kg/m$^2$ (stratified analyses): | | | |
| < 20.0 kg/m$^2$ | 0.64 (0.38 to 1.07) | 0.70 (0.42 to 1.15) | 0.67 (0.39 to 1.16) |
| 20.0-24.9 kg/m$^2$ | 0.96 (0.88 to 1.04) | 0.97 (0.89 to 1.05) | 0.95 (0.87 to 1.05) |
| 25.0-29.9 kg/m$^2$ | 1.01 (0.93 to 1.09) | 1.01 (0.94 to 1.10) | 1.04 (0.95 to 1.14) |
| ≥ 30.0 kg/m$^2$ | 1.23 (1.06 to 1.43) | 1.26 (1.08 to 1.48) | 1.24 (1.05 to 1.46) |
| BMI with lowest subfertility odds | 25.2 kg/m$^2$ | 25.0 kg/m$^2$ | 25.2 kg/m$^2$ |

*ART*: assisted reproductive technology; *MR*: Mendelian randomization; *OR*: odds ratio.

### SUPPLEMENTAL REFERENCES


1. Helgeland Ø, Vaudel M, Juliusson PB, Lingaas Holmen O, Juodakis J, Bacelis J, et al. Genome-wide association study reveals dynamic role of genetic variation in infant and early childhood growth. Nat Commun. 2019;10(1):4448. doi: 10.1038/s41467-019-12308-0.
2. Sole-Navais P, Bacelis J, Helgeland Ø, Modzelewska D, Vaudel M, Flatley C, et al. Autozygosity mapping and time-to-spontaneous delivery in Norwegian parent-offspring trios. Hum Mol Genet. 2021;29(23):3845-58. doi: 10.1093/hmg/ddaa255.
3. Chang CC, Chow CC, Tellier LC, Vattikuti S, Purcell SM, Lee JJ. Second-generation PLINK: rising to the challenge of larger and richer datasets. Gigascience. 2015;4:7. doi: 10.1186/s13742-015-0047-8.
4. Mägi R, Morris AP. GWAMA: software for genome-wide association meta-analysis. BMC Bioinformatics. 2010;11:288. doi: 10.1186/1471-2105-11-288.
